# A Longitudinal Normative Dataset and Protocol for Speech and Language Biomarker Research

**DOI:** 10.1101/2021.08.16.21262125

**Authors:** James W. Schwoebel, Joel Schwartz, Lindsay A. Warrenburg, Roland Brown, Ashi Awasthi, Austin New, Monroe Butler, Mark Moss, Eleftheria K. Pissadaki

## Abstract

Although speech and language biomarker (SLB) research studies have shown methodological and clinical promise, some common limitations of these studies include small sample sizes, limited longitudinal data, and a lack of a standardized survey protocol. Here, we introduce the Voiceome Protocol and the corresponding Voiceome Dataset as standards which can be utilized and adapted by other SLB researchers. The Voiceome Protocol includes 12 types of voice tasks, along with health and demographic questions that have been shown to affect speech. The longitudinal Voiceome Dataset consisted of the Voiceome Protocol survey taken on (up to) four occasions, each separated by roughly three weeks (22.80 +/- 20.91 days). Of 6,650 total participants, 1,382 completed at least two Voiceome surveys. The results of the Voiceome Dataset are largely consistent with results from standard clinical literature, suggesting that the Voiceome Study is a high-fidelity, normative dataset and scalable protocol that can be used to advance SLB research.

## Main Manuscript

Speech and language biomarkers (SLBs) have emerged as a medium to detect changes in cognition and health. Individuals with mild cognitive impairment can be distinguished from healthy controls (Bertola et al., 2014) from a combination of speech features from multiple language tasks (Eyigoz, Mathur, Santamaria, Cecchi, & Naylor, 2020) and by employing machine learning architectures to train a series of cascaded classifiers (Fraser et al., 2019). Bedi and colleagues furthermore demonstrated that it is possible to train a classification model with 100% accuracy to predict psychosis onset in at-risk youth with three speech features—semantic coherence, maximum phrase length, and use of determiners—derived from a free speech task, outperforming classification from clinical interviews (Bedi et al., 2015). Custom engineered speech landmark features—such as *glottis*, a sustained vibration of the vocal folds starts and ends—have been used to characterize depression symptoms (Huang, Epps, & Joachim, 2019). Recent research is also consistent with the idea that speech-based machine learning models can be used to detect COVID-19 status (Bagad et al., 2020). In the neurology and motor coordination domain, machine learning models have been shown to discriminate Parkinson’s disease patients from controls with an accuracy of 85%, which exceeds the average clinical diagnosis accuracy of non-experts (73.8%) and average accuracy of movement disorder specialists (79.6% without follow-up, 83.9% after follow-up; Wroge et al., 2018). These studies, among others, suggest the promise of using SLBs to detect health changes over time.

Some datasets have served as SLB benchmarks, with which other clinical studies can compare speech metrics. These benchmark datasets exist for number of health conditions, including Alzheimer’s disease, dementia, respiratory conditions, Parkinson’s disease, and clinical depression (Table 1). At times, these datasets have been used for public machine learning challenges, such as the Interspeech DiCOVA 2021 challenge and the ADReSSo Challenge. The goal of these public challenges is two-fold: (1) to spur translation of new featurization and modeling techniques, and (2) to develop a definition of state-of-the-art model performance.

**Table 1.**
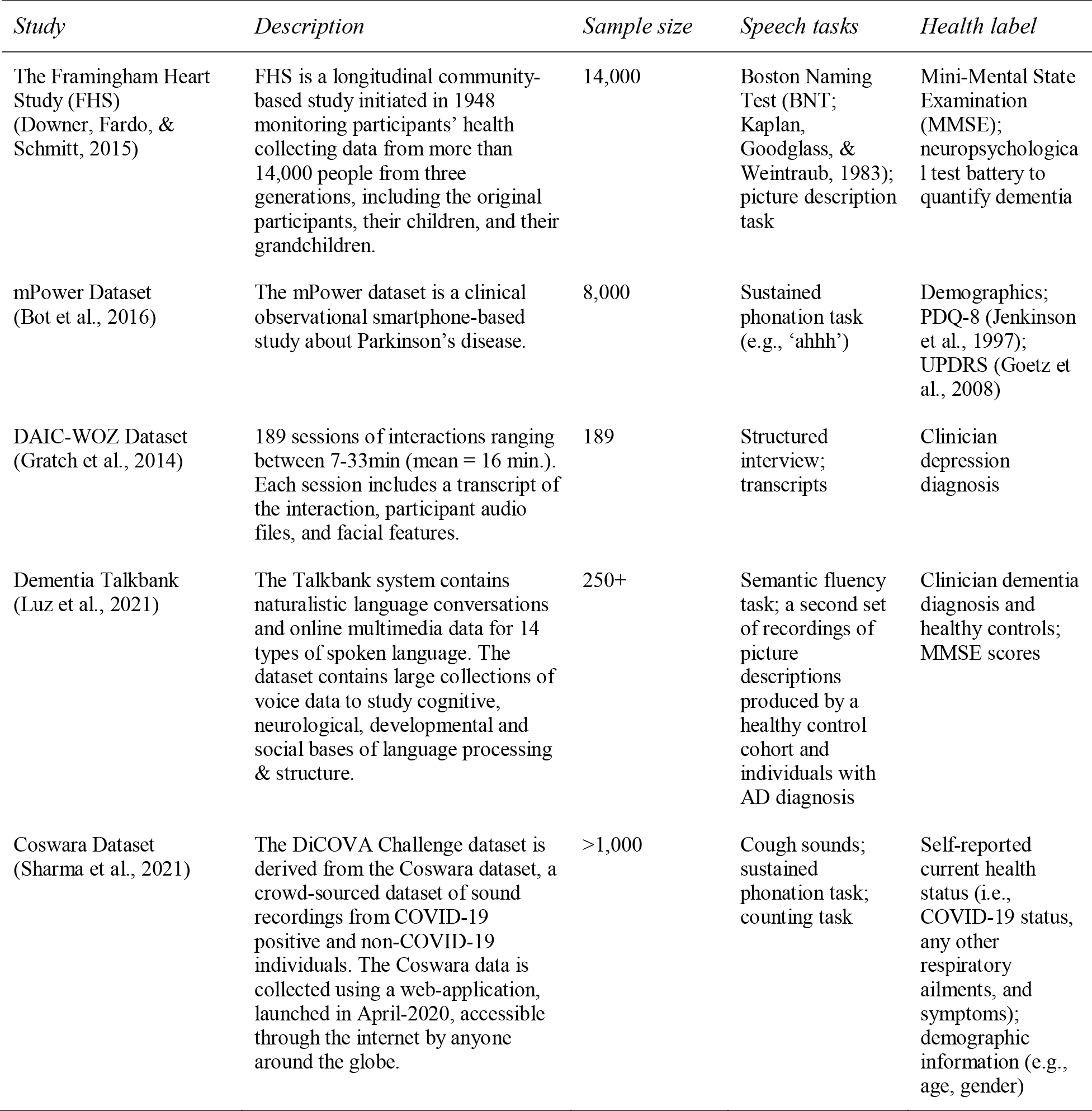
Overview of standard Speech and Language vocal Biomarker (SLB) datasets

| *Study* | *Description* | *Sample size* | *Speech tasks* | *Health label* |
| --- | --- | --- | --- | --- |
| The Framingham Heart Study (FHS) (Downer, Fardo, & Schmitt, 2015) | FHS is a longitudinal community-based study initiated in 1948 monitoring participants' health collecting data from more than 14,000 people from three generations, including the original participants, their children, and their grandchildren. | 14,000 | Boston Naming Test (BNT; Kaplan, Goodglass, & Weintraub, 1983); picture description task | Mini-Mental State Examination (MMSE); neuropsychological test battery to quantify dementia |
| mPower Dataset (Bot et al., 2016) | The mPower dataset is a clinical observational smartphone-based study about Parkinson's disease. | 8,000 | Sustained phonation task (e.g., 'ahhh') | Demographics; PDQ-8 (Jenkinson et al., 1997); UPDRS (Goetz et al., 2008) |
| DAIC-WOZ Dataset (Gratch et al., 2014) | 189 sessions of interactions ranging between 7-33min (mean = 16 min.). Each session includes a transcript of the interaction, participant audio files, and facial features. | 189 | Structured interview; transcripts | Clinician depression diagnosis |
| Dementia Talkbank (Luz et al., 2021) | The Talkbank system contains naturalistic language conversations and online multimedia data for 14 types of spoken language. The dataset contains large collections of voice data to study cognitive, neurological, developmental and social bases of language processing & structure. | 250+ | Semantic fluency task; a second set of recordings of picture descriptions produced by a healthy control cohort and individuals with AD diagnosis | Clinician dementia diagnosis and healthy controls; MMSE scores |
| Coswara Dataset (Sharma et al., 2021) | The DiCOVA Challenge dataset is derived from the Coswara dataset, a crowd-sourced dataset of sound recordings from COVID-19 positive and non-COVID-19 individuals. The Coswara data is collected using a web-application, launched in April-2020, accessible through the internet by anyone around the globe. | >1,000 | Cough sounds; sustained phonation task; counting task | Self-reported current health status (i.e., COVID-19 status, any other respiratory ailments, and symptoms); demographic information (e.g., age, gender) |

There are several limitations with these public challenges. When viewed holistically, most of these standard datasets might be seen as biased with regards to a binary classification of health condition versus a control group. Additionally, most public challenge datasets contain confounding factors with regard to health condition detection (e.g., age, gender), contain relatively small sample sizes (*n* < 500 individuals), and have relatively few speech tasks or prompts (usually less than five). Furthermore, the datasets tend to require substantial data cleaning before they can be interpreted by the challenge participants. Finally, these datasets may not be typically representative of the standard United States population (e.g., by age, gender, and location; de la Fuente Garcia, Craig, & Luz, 2020) and demographic variables have reportedly been established as risk factors (Mielke, Vemuri, & Rocca, 2014). These limitations result in most challenge solutions being overfit to a specific context. The solutions therefore usually lack the ability to be generalized beyond the challenge dataset or scaled to additional contexts, participants, and studies.

To address these shortcomings, review papers have proposed best practices for SLB-related research (de la Fuente Garcia, Craig, & Luz, 2020; Low, Bentley, & Ghosh, 2020; Robin et al., 2020). These guidelines suggest that when creating new datasets, it is important to do the following things:

1. Report health comorbidities
2. Focus on detecting symptoms or specific problems instead of entire health conditions
3. Consider additional confounds when selecting control groups
4. Compare multiple operationalizations of health assessments (e.g., self-report vs. clinical diagnosis)
5. Use power analysis to determine sample size for null_Jhypothesis testing
6. Include multiple speech tasks and prompts for cross-sectional and longitudinal data
7. Use one microphone per speaker in recorded interviews
8. Use standard acoustic featurization techniques (e.g., Allie repository, Schwoebel, 2020; GeMAPS, Eyben et al., 2016)
9. Check the accuracy and reliability of data processing and computed measures (e.g., test-retest reliability, comparing speech measures to reference standards)

Despite some progress, there remain few normative datasets that can be used to benchmark the performance of SLBs across a range of speech tasks, microphone types, featurization methods, and modeling techniques that follow standard best practices (Low, Bentley, & Ghosh, 2020). As most SLB study paradigms were designed to investigate a specific health condition, they therefore tend to consist of a small number of focused speech tasks, as well.

The Voiceome Protocol and corresponding Voiceome Dataset were created in response to the above limitations. The Voiceome Protocol employs a comprehensive battery of twelve types of speech tasks that can be applied across a range of health conditions. The primary goal of the Voiceome Protocol is to provide an easy-to-use template for future SLB-related research studies with regards to study design and protocol.

The Voiceome Dataset utilized the main survey from the Voiceome Protocol in a study of more than six thousand participants. The study was longitudinal in design, where participants were asked to complete the Voiceome survey on four occasions, each occurrence separated by roughly three weeks. The main goal of the Voiceome Dataset is to provide voice metric standards for a representative population sample of the United States, with which other SLB researchers can compare their study results.

Taken together, the Voiceome Protocol and Study aim to do the following:

1. Establish a longitudinal reference protocol with a wide variety of speech tasks
2. Define quality standards and reference features for novel and typical SLB-related tasks
3. Identify confounding factors related to SLB-related research studies
4. Demonstrate the ability to conduct large scale, decentralized clinical studies for SLBs using SurveyLex, a tool to create and clone web-based voice surveys in less than 1 minute (https://www.surveylex.com).

## Results

### Participants

All study materials and procedures were approved by the Western Institutional Review Board (protocol #20170781). Participant enrollment was open to individuals that were U.S. residents 18 years of age or older who self-reported feeling comfortable reading and writing in English. All participants were required to have access to a device with a microphone and with an internet connection. Various methods were used to recruit participants, including Google Ads, Facebook Ads, Amazon Mechanical Turk (mTurk), email newsletters (e.g., through NAMI), tailored LinkedIn messages, and through personal outreach. Due to the most effective cost per acquisition, Amazon Mechanical Turk was used predominantly for recruiting participants. The Methods section details recruitment methods for the Voiceome Dataset (Figure 7) for details survey completion and attrition.

Overall, participant demographics were representative of the United States population (Table 2 and Figure D.2). Participants were 34.6% male and 64.3% female, had an average age of 33.95 years (SD = 11.90), and an average BMI of 27.20 (SD = 7.24). 65.7% of participants were White, 10.0% were Black or African American, 8.9% were Asian or Asian American, and 15.4% reported being another race or ethnicity. Roughly 96% of participants were United States residents and 91.8% reported speaking English as their first language. With regard to health conditions, participants were primarily non-depressed (PHQ-9: M = 4.50, SD = 3.71), non-anxious (GAD-7: M = 4.05, SD = 3.42), and reported feeling reasonably well (*On a scale of 1-10, how well do you feel right now*, anchored at 1 ‘not at all well’ and 10 ‘extremely well’: M = 7.65, SD = 1.64). 10.1% of participants reported being diagnosed with clinical depression and 2.02% of subjects took Zoloft to treat their depression or anxiety symptoms.

**Table 2.**
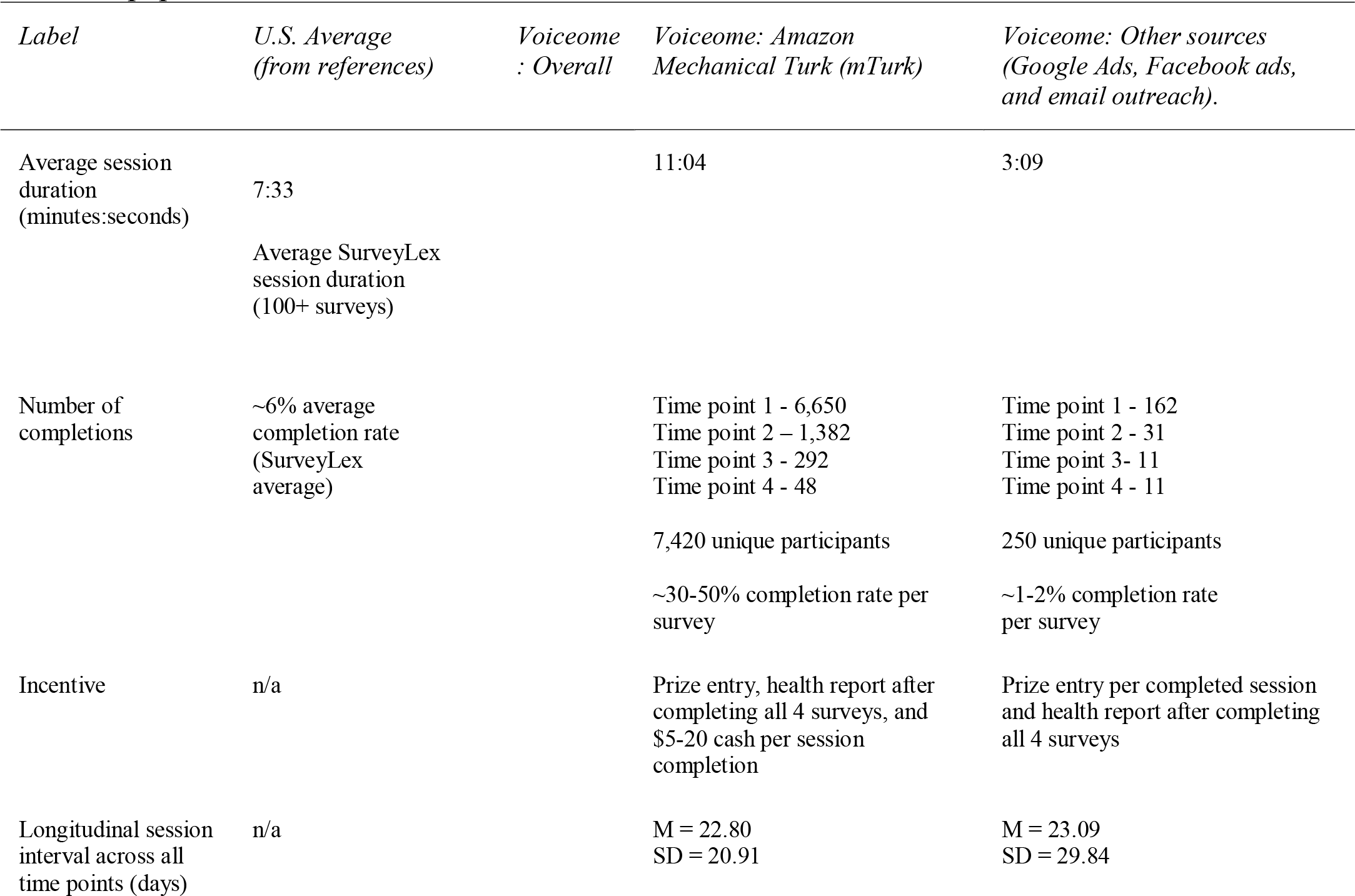
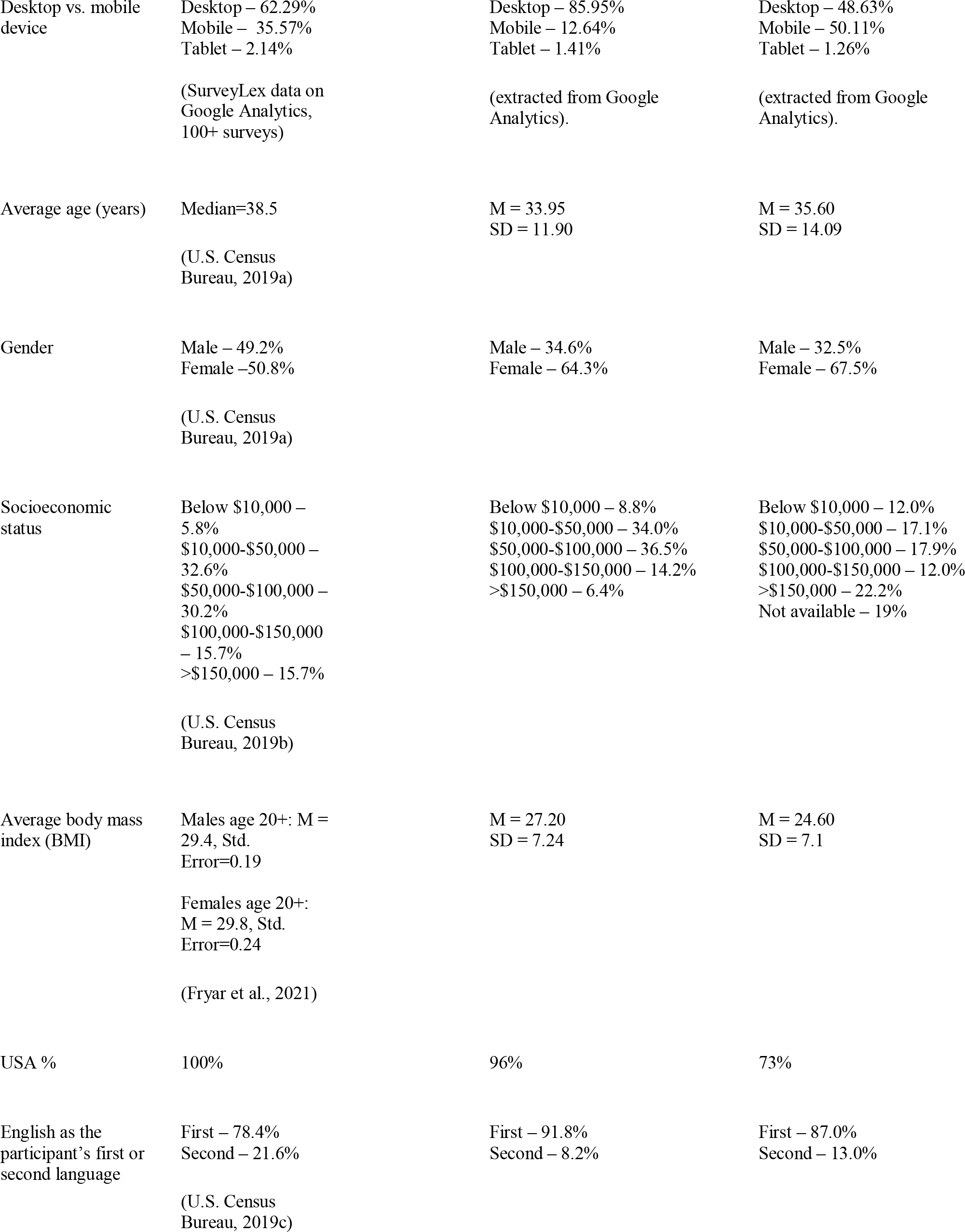
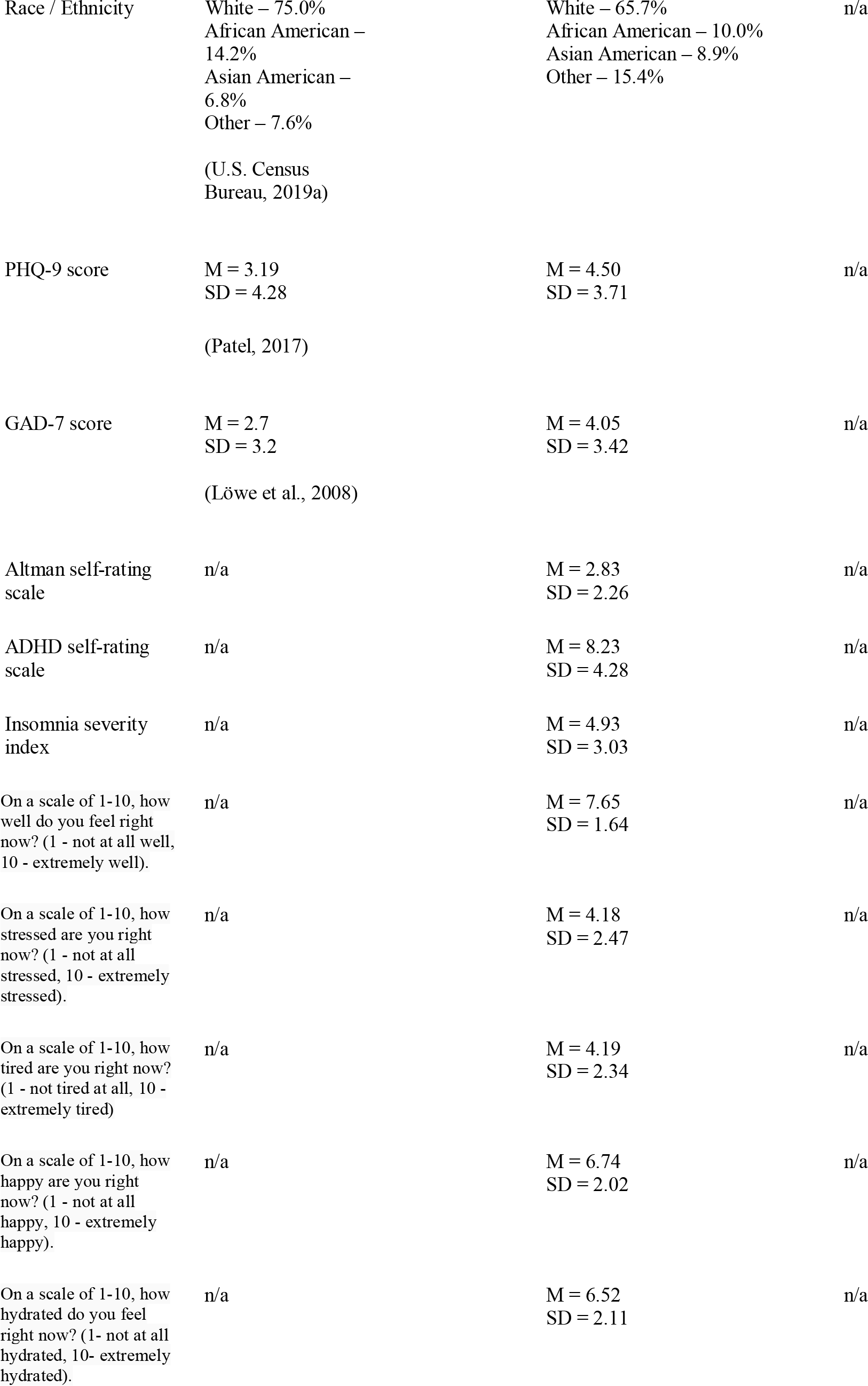
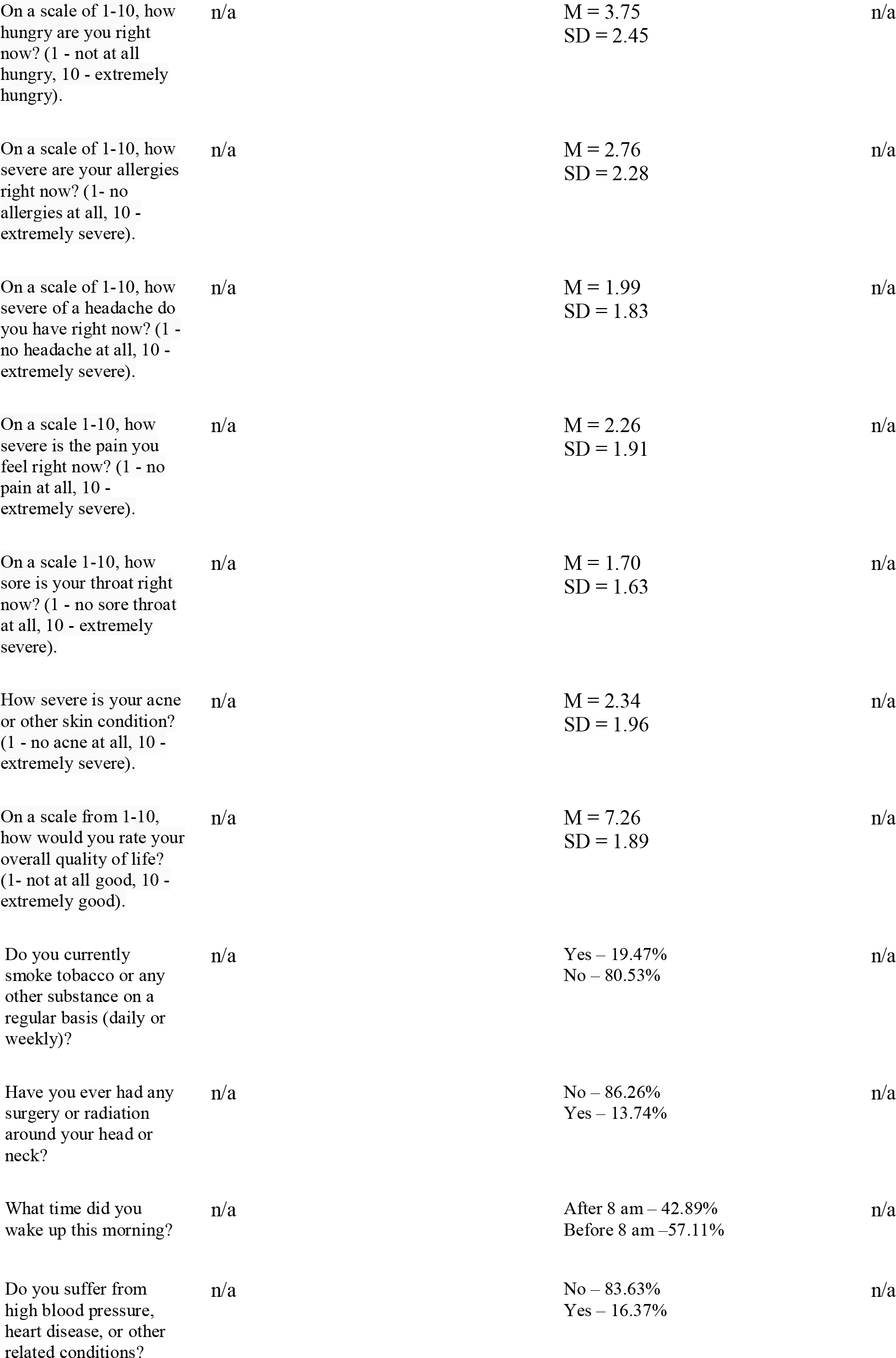
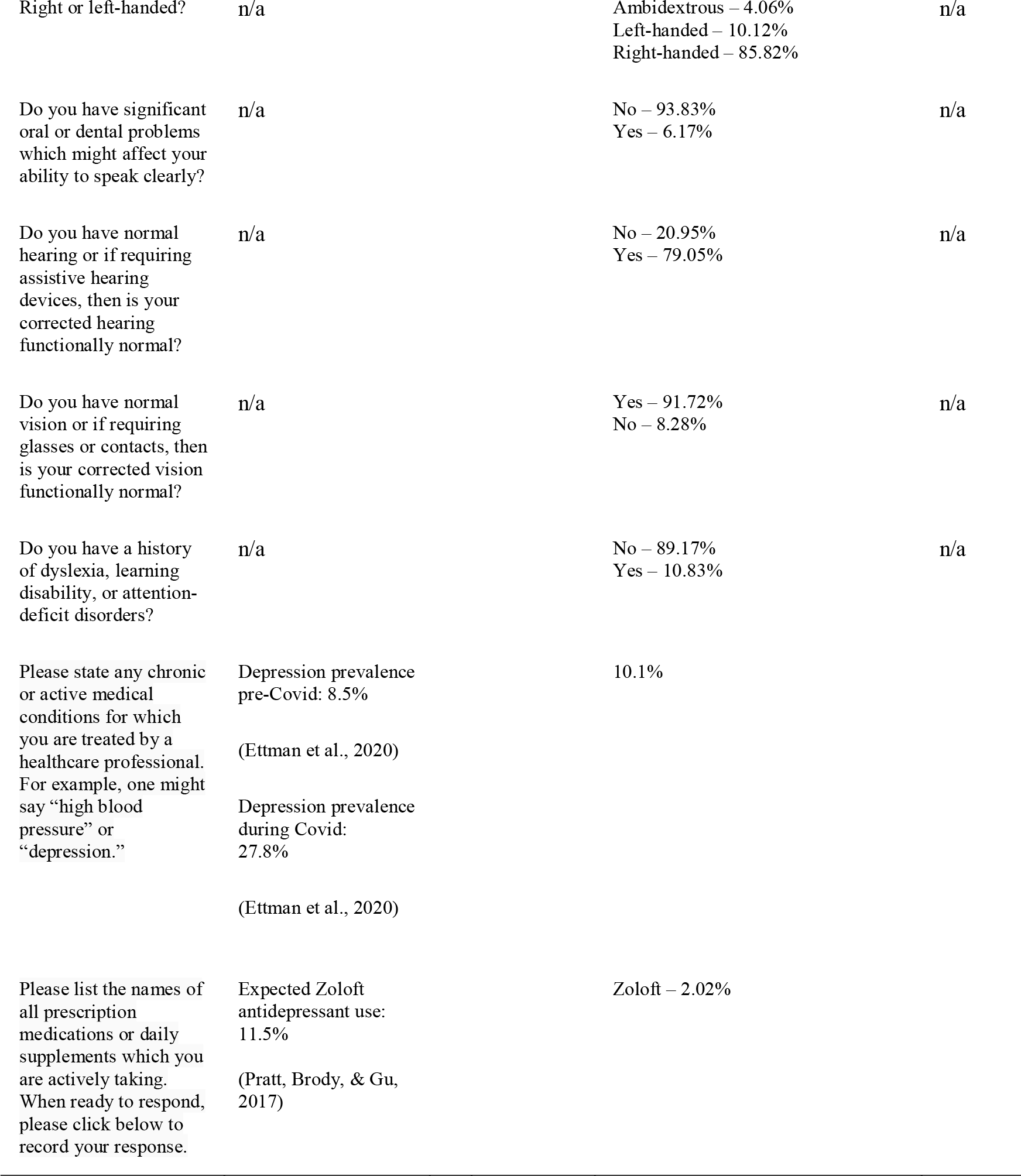
Voiceome Dataset participant enrollment and demographic information, compared to the U.S. population.

| *Label* | *U.S. Average*<br>*(from references)* | *Voiceome*<br>*: Overall* | *Voiceome: Amazon*<br>*Mechanical Turk (mTurk)* | *Voiceome: Other sources*<br>*(Google Ads, Facebook ads,*<br>*and email outreach).* |
| --- | --- | --- | --- | --- |
| Average session duration<br>(minutes:seconds) | 7:33<br>Average SurveyLex session duration<br>(100+ surveys) |  | 11:04 | 3:09 |
| Number of completions | ~6% average completion rate<br>(SurveyLex average) |  | Time point 1 - 6,650<br>Time point 2 - 1,382<br>Time point 3 - 292<br>Time point 4 - 48<br>7,420 unique participants<br>~30-50% completion rate per survey | Time point 1 - 162<br>Time point 2 - 31<br>Time point 3 - 11<br>Time point 4 - 11<br>250 unique participants<br>~1-2% completion rate per survey |
| Incentive | n/a | | Prize entry, health report after completing all 4 surveys, and \$5-20 cash per session completion | Prize entry per completed session and health report after completing all 4 surveys |
| Longitudinal session interval across all time points (days) | n/a | | $M = 22.80$<br>$SD = 20.91$ | $M = 23.09$<br>$SD = 29.84$ |

|  |  |  |  |
| --- | --- | --- | --- |
| Desktop vs. mobile device | Desktop – 62.29%<br>Mobile – 35.57%<br>Tablet – 2.14%<br>(SurveyLex data on Google Analytics, 100+ surveys) | Desktop – 85.95%<br>Mobile – 12.64%<br>Tablet – 1.41%<br>(extracted from Google Analytics). | Desktop – 48.63%<br>Mobile – 50.11%<br>Tablet – 1.26%<br>(extracted from Google Analytics). |
| Average age (years) | Median=38.5<br>(U.S. Census Bureau, 2019a) | M = 33.95<br>SD = 11.90 | M = 35.60<br>SD = 14.09 |
| Gender | Male – 49.2%<br>Female – 50.8%<br>(U.S. Census Bureau, 2019a) | Male – 34.6%<br>Female – 64.3% | Male – 32.5%<br>Female – 67.5% |
| Socioeconomic status | Below \$10,000 – 5.8%<br>\$10,000-\$50,000 – 32.6%<br>\$50,000-\$100,000 – 30.2%<br>\$100,000-\$150,000 – 15.7%<br>>\$150,000 – 15.7%<br>(U.S. Census Bureau, 2019b) | Below \$10,000 – 8.8%<br>\$10,000-\$50,000 – 34.0%<br>\$50,000-\$100,000 – 36.5%<br>\$100,000-\$150,000 – 14.2%<br>>\$150,000 – 6.4% | Below \$10,000 – 12.0%<br>\$10,000-\$50,000 – 17.1%<br>\$50,000-\$100,000 – 17.9%<br>\$100,000-\$150,000 – 12.0%<br>>\$150,000 – 22.2%<br>Not available – 19% |
| Average body mass index (BMI) | Males age 20+: M = 29.4, Std. Error=0.19<br>Females age 20+: M = 29.8, Std. Error=0.24<br>(Fryar et al., 2021) | M = 27.20<br>SD = 7.24 | M = 24.60<br>SD = 7.1 |
| USA % | 100% | 96% | 73% |
| English as the participant's first or second language | First – 78.4%<br>Second – 21.6%<br>(U.S. Census Bureau, 2019c) | First – 91.8%<br>Second – 8.2% | First – 87.0%<br>Second – 13.0% |

|  |  |  |  |
| --- | --- | --- | --- |
| Race / Ethnicity | White – 75.0%<br>African American – 14.2%<br>Asian American – 6.8%<br>Other – 7.6%<br>(U.S. Census Bureau, 2019a) | White – 65.7%<br>African American – 10.0%<br>Asian American – 8.9%<br>Other – 15.4% | n/a |
| PHQ-9 score | M = 3.19<br>SD = 4.28<br>(Patel, 2017) | M = 4.50<br>SD = 3.71 | n/a |
| GAD-7 score | M = 2.7<br>SD = 3.2<br>(Löwe et al., 2008) | M = 4.05<br>SD = 3.42 | n/a |
| Altman self-rating scale | n/a | M = 2.83<br>SD = 2.26 | n/a |
| ADHD self-rating scale | n/a | M = 8.23<br>SD = 4.28 | n/a |
| Insomnia severity index | n/a | M = 4.93<br>SD = 3.03 | n/a |
| On a scale of 1-10, how well do you feel right now? (1 - not at all well, 10 - extremely well). | n/a | M = 7.65<br>SD = 1.64 | n/a |
| On a scale of 1-10, how stressed are you right now? (1 - not at all stressed, 10 - extremely stressed). | n/a | M = 4.18<br>SD = 2.47 | n/a |
| On a scale of 1-10, how tired are you right now? (1 - not tired at all, 10 - extremely tired) | n/a | M = 4.19<br>SD = 2.34 | n/a |
| On a scale of 1-10, how happy are you right now? (1 - not at all happy, 10 - extremely happy). | n/a | M = 6.74<br>SD = 2.02 | n/a |
| On a scale of 1-10, how hydrated do you feel right now? (1- not at all hydrated, 10- extremely hydrated). | n/a | M = 6.52<br>SD = 2.11 | n/a |

|  |  |  |  |
| --- | --- | --- | --- |
| On a scale of 1-10, how hungry are you right now? (1 - not at all hungry, 10 - extremely hungry). | n/a | M = 3.75<br>SD = 2.45 | n/a |
| On a scale of 1-10, how severe are your allergies right now? (1- no allergies at all, 10 - extremely severe). | n/a | M = 2.76<br>SD = 2.28 | n/a |
| On a scale of 1-10, how severe of a headache do you have right now? (1 - no headache at all, 10 - extremely severe). | n/a | M = 1.99<br>SD = 1.83 | n/a |
| On a scale 1-10, how severe is the pain you feel right now? (1 - no pain at all, 10 - extremely severe). | n/a | M = 2.26<br>SD = 1.91 | n/a |
| On a scale 1-10, how sore is your throat right now? (1 - no sore throat at all, 10 - extremely severe). | n/a | M = 1.70<br>SD = 1.63 | n/a |
| How severe is your acne or other skin condition? (1 - no acne at all, 10 - extremely severe). | n/a | M = 2.34<br>SD = 1.96 | n/a |
| On a scale from 1-10, how would you rate your overall quality of life? (1- not at all good, 10 - extremely good). | n/a | M = 7.26<br>SD = 1.89 | n/a |
| Do you currently smoke tobacco or any other substance on a regular basis (daily or weekly)? | n/a | Yes – 19.47%<br>No – 80.53% | n/a |
| Have you ever had any surgery or radiation around your head or neck? | n/a | No – 86.26%<br>Yes – 13.74% | n/a |
| What time did you wake up this morning? | n/a | After 8 am – 42.89%<br>Before 8 am – 57.11% | n/a |
| Do you suffer from high blood pressure, heart disease, or other related conditions? | n/a | No – 83.63%<br>Yes – 16.37% | n/a |

|  |  |  |  |
| --- | --- | --- | --- |
| Right or left-handed? | n/a | Ambidextrous – 4.06%<br>Left-handed – 10.12%<br>Right-handed – 85.82% | n/a |
| Do you have significant oral or dental problems which might affect your ability to speak clearly? | n/a | No – 93.83%<br>Yes – 6.17% | n/a |
| Do you have normal hearing or if requiring assistive hearing devices, then is your corrected hearing functionally normal? | n/a | No – 20.95%<br>Yes – 79.05% | n/a |
| Do you have normal vision or if requiring glasses or contacts, then is your corrected vision functionally normal? | n/a | Yes – 91.72%<br>No – 8.28% | n/a |
| Do you have a history of dyslexia, learning disability, or attention-deficit disorders? | n/a | No – 89.17%<br>Yes – 10.83% | n/a |
| Please state any chronic or active medical conditions for which you are treated by a healthcare professional. For example, one might say “high blood pressure” or “depression.” | Depression prevalence pre-Covid: 8.5%<br>(Ettman et al., 2020)<br>Depression prevalence during Covid: 27.8%<br>(Ettman et al., 2020) | 10.1% |  |
| Please list the names of all prescription medications or daily supplements which you are actively taking. When ready to respond, please click below to record your response. | Expected Zoloft antidepressant use: 11.5%<br>(Pratt, Brody, & Gu, 2017) | Zoloft – 2.02% |  |

### Speech Tasks

Voiceome Dataset participants completed twelve types of speech tasks (Table 3), many of which mirror speech tasks that have been used in the clinical literature (e.g., Kaploun et al., 2011; Mahler, 2012; Maslan et al., 2011; Opasso, Barreto, & Ortiz, 2016; Patel et al., 2013; Vaughan et al., 2018). As the overall demographics and depression prevalence of study participants are similar in the Voiceome Dataset and other peer-reviewed clinical studies, it is illuminating to compare performance metrics of these speech tasks between the Voiceome and previous research. In general, the results of Voiceome participants for each task mirrored what was expected from peer-reviewed clinical norms. The Voiceome Dataset also reveals performance metrics for speech tasks previously untested in the clinical literature (Table 3). The following analyses refer to 2,465 participants who completed Survey A between March 2019 and May 2020. Table 4 documents the Voiceome Dataset performance metrics of all twelve speech tasks for all 2,465 participants, as well as by gender and age cohorts.

**Table 3.**
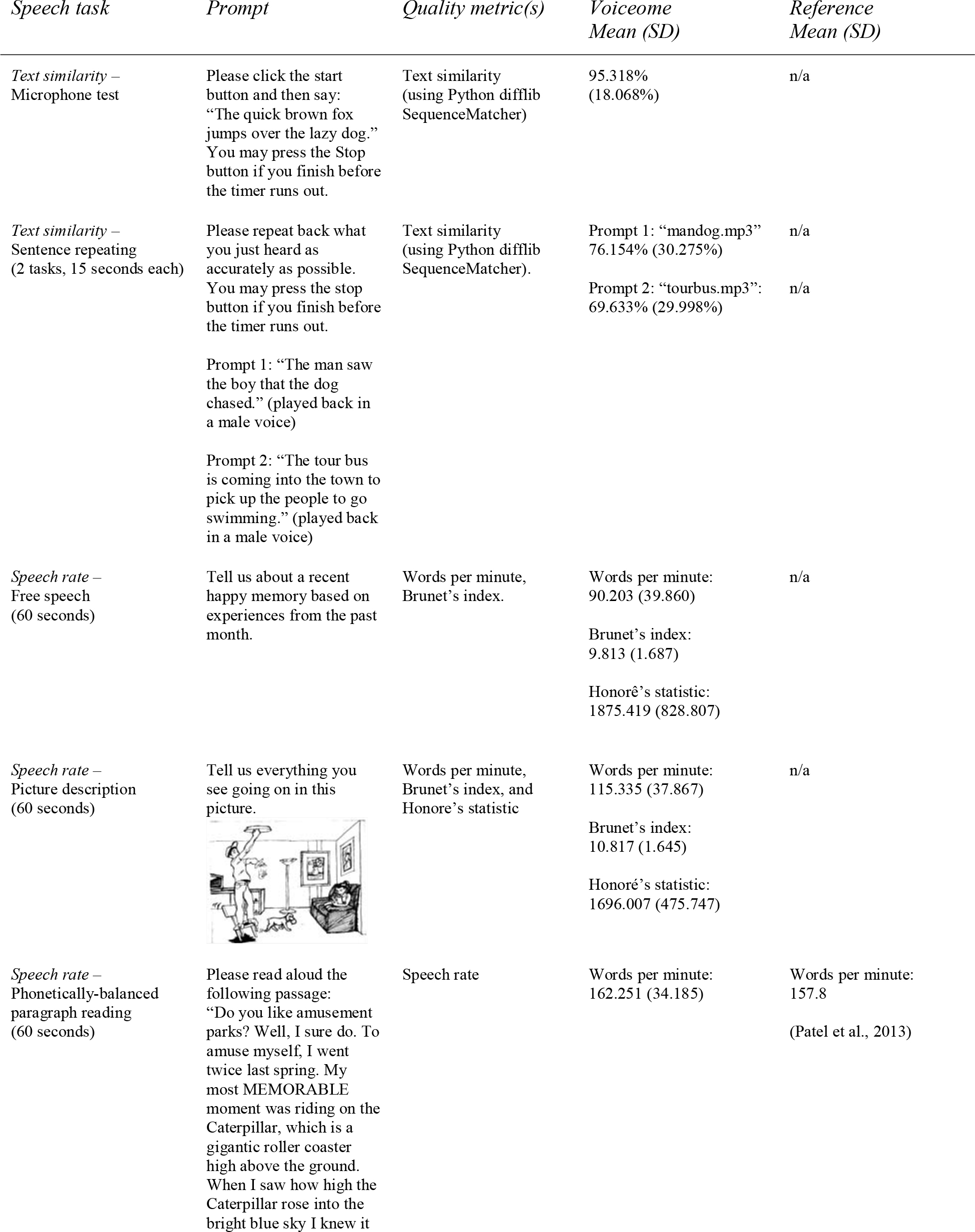
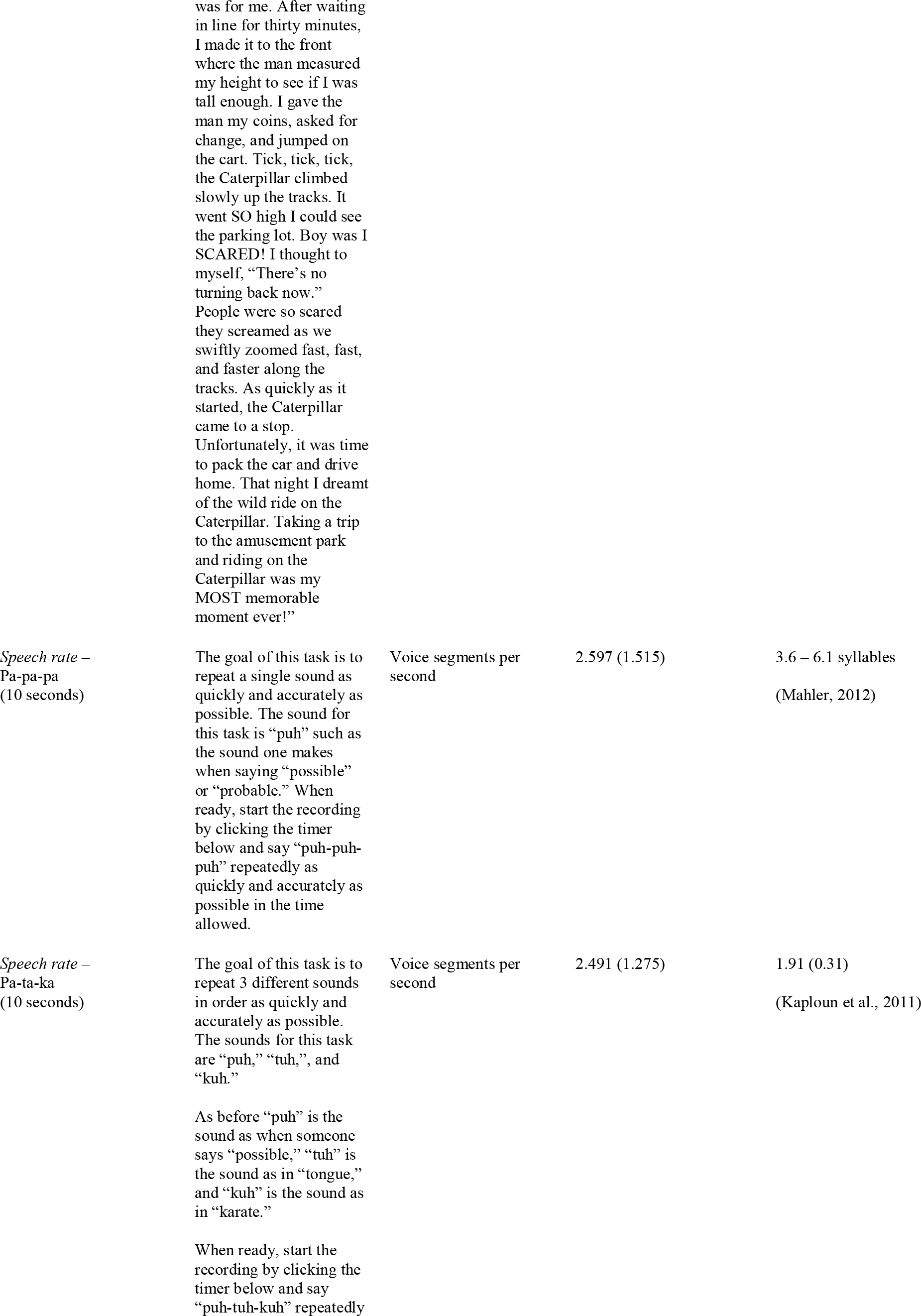
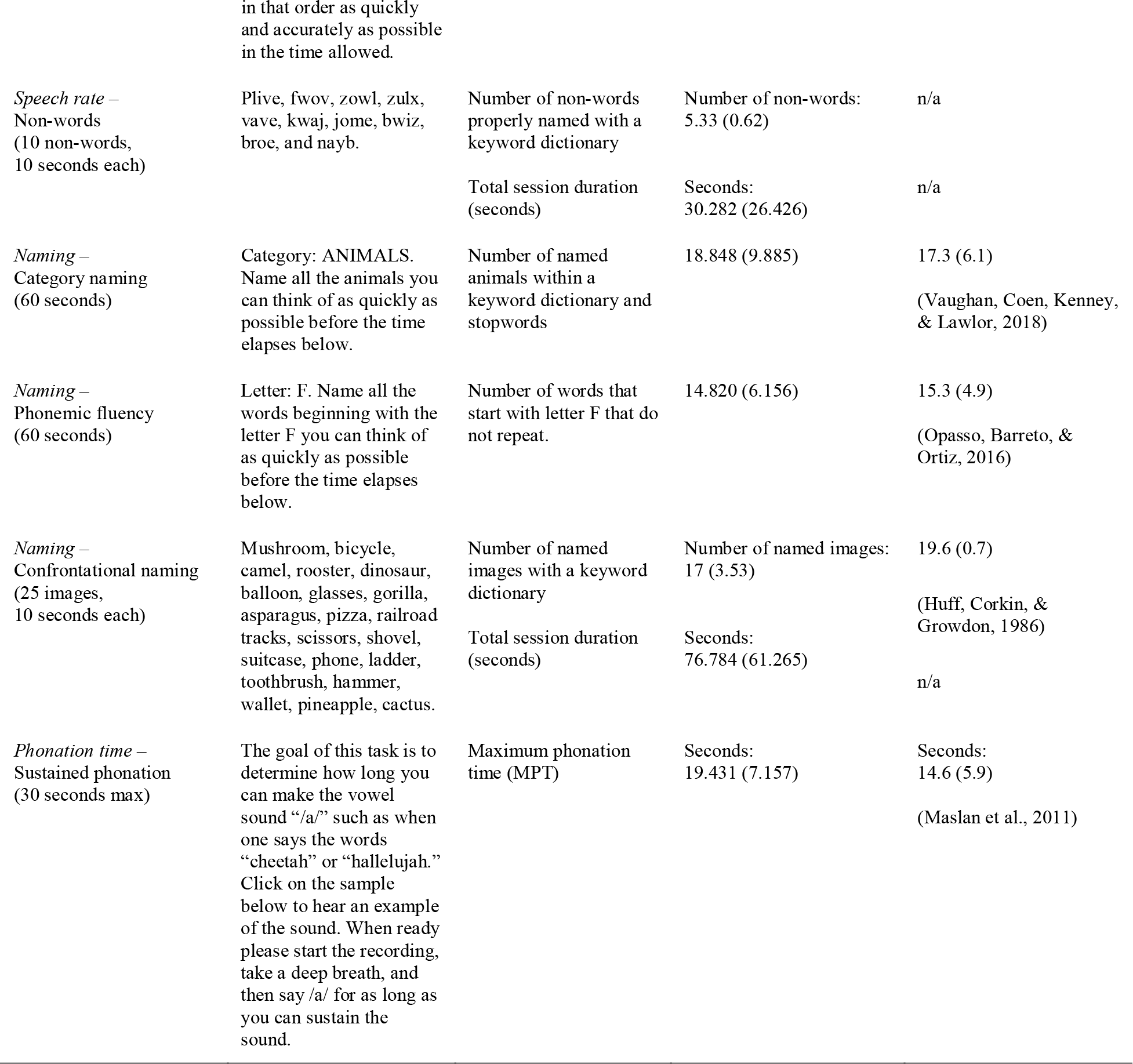
Comparison of Voiceome speech tasks results and reference results. For more details on how these values vary by age and gender, please see the Supplemental Materials and the Voiceome GitHub page: https://github.com/jim-schwoebel/voiceome.

**Table 4.**
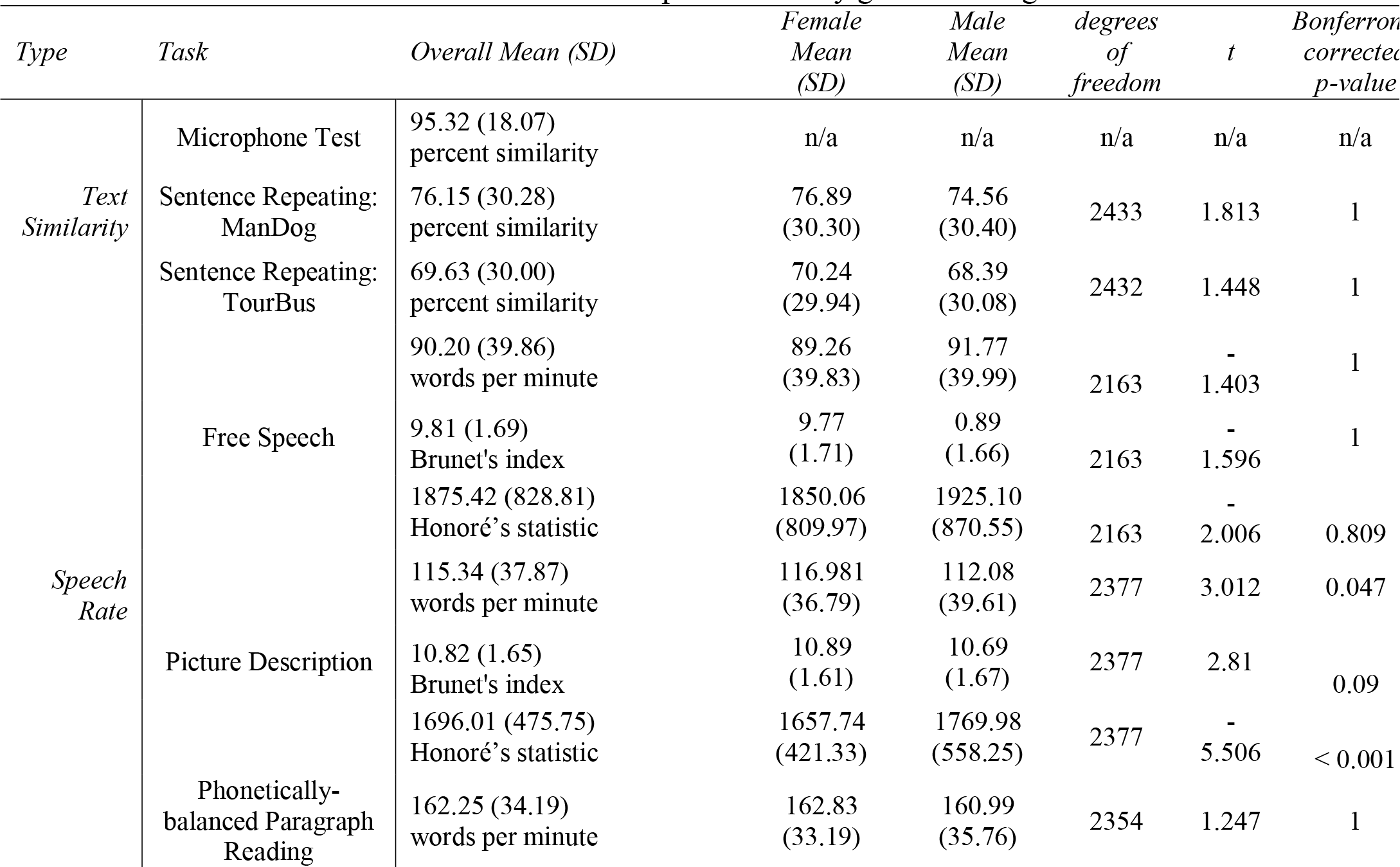
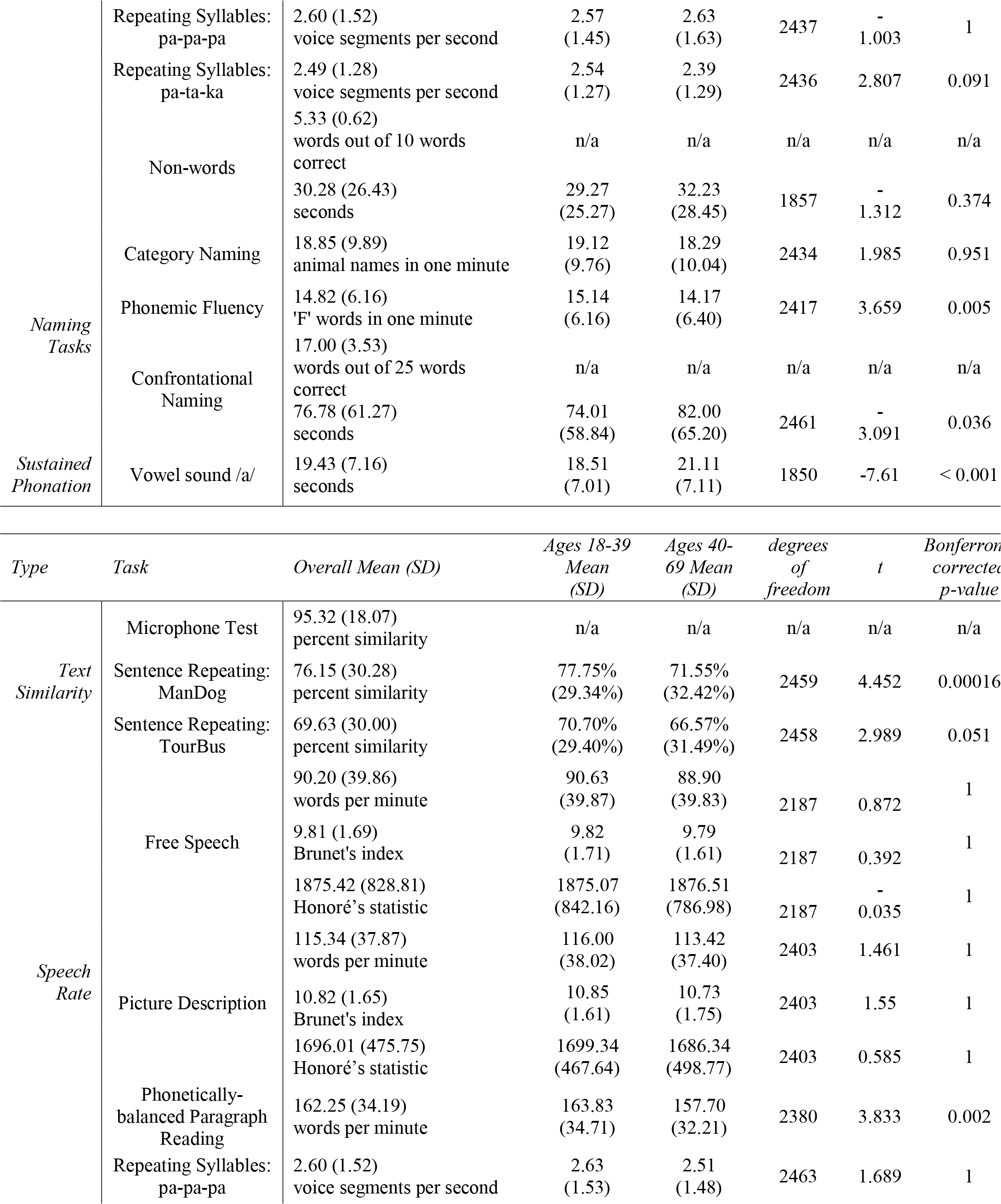
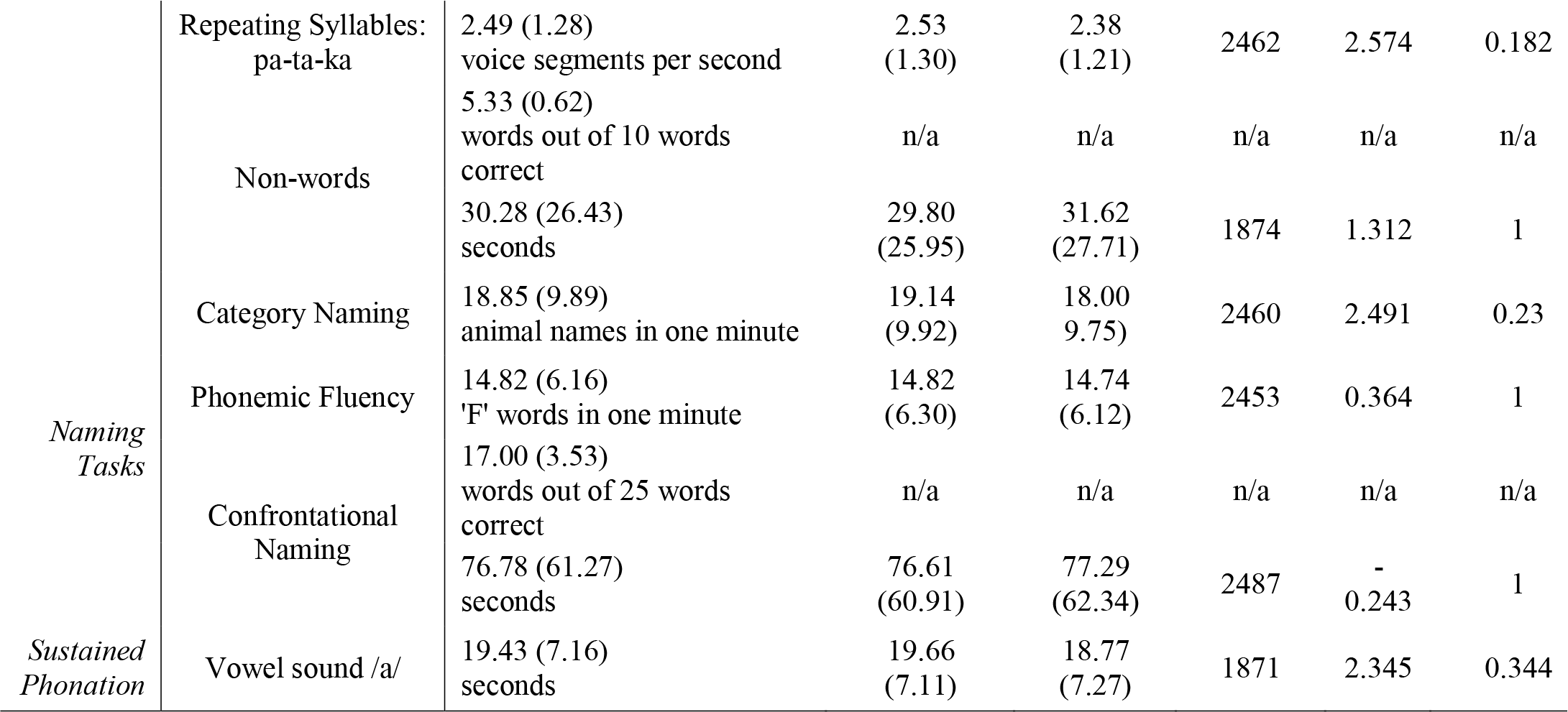
Voiceome Dataset results for all twelve speech tasks by gender and age bracket.

Figure 1 shows a t-SNE plot summarizing the independence of all speech tasks from Survey A. The t-Distributed Stochastic Neighbor Embedding (t-SNE) method enables highly dimensional data to be visualized in a two-dimensional space. Visualizing Voiceome speech data using t-SNE enables us to check how many distinct dimensions exist in the data, as well as which tasks may provide overlapping information. Figure 1 shows that most speech tasks clustered independently, indicating that they provide non-overlapping information. For example, the picture description task, the animal naming task, and the Caterpillar naming task each have a distinctive cluster. There were only two cases where speech tasks produced overlapping clusters. First, the pa-pa-pa and pa-ta-ka tasks overlapped, possibly because of their overlapping elicitations (‘pa’). Second, free speech tasks with similar semantic content—such as the listing of medications and listing of health diagnosis—tended to cluster together. The medication and diagnosis prompt elicited responses that commonly began with similar wording, such as ‘I do not have any…’. Overall, the t-SNE plot shows that the twelve different speech tasks in the Voiceome Dataset each provided unique information about the speaker.

**Figure 1.**
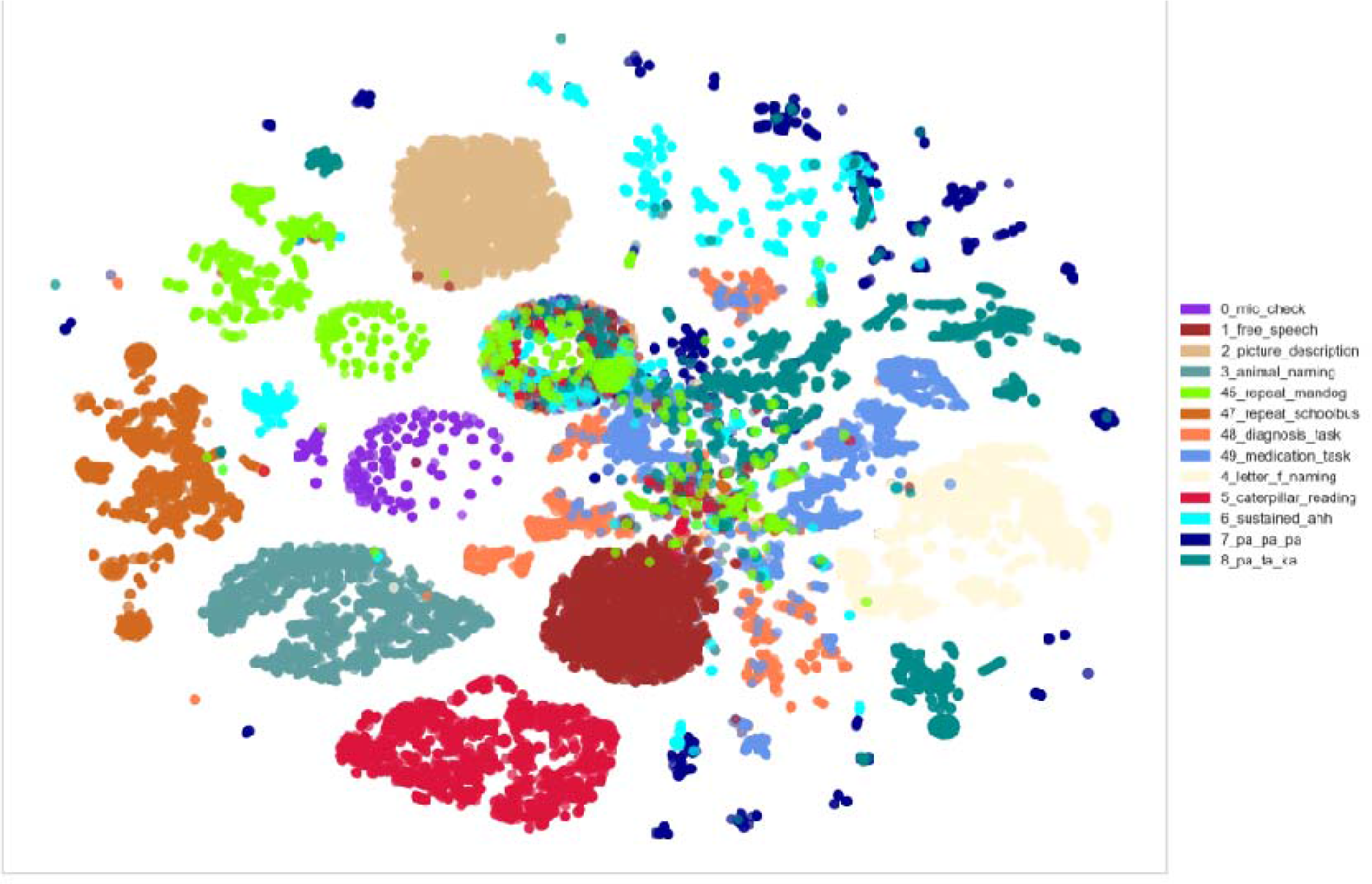
t-SNE plot from Survey A responses for all speech tasks. The figure demonstrates that most speech tasks clustered independently, except for tasks with overlapping elicitations (e.g., ‘pa-pa-pa’ and ‘pa-ta-ka’ tasks) or tasks that yield similar transcripts (e.g., medication and diagnosis prompts, whose responses commonly begin with similar wording, such as ‘I do not have any…’).

### Text similarity

#### Microphone test

In order to make sure participants’ microphones were working, they were asked to repeat the phrase, *The quick brown fox jumps over the lazy dog*. The performance metric to evaluate this task is to compare the similarity between the words the participants say and the reference sentence. The Voiceome participants had an overall average of 95.32% similarity (SD = 18.07%) relative to the reference sentence.

#### Sentence repeating

Voiceome participants were asked to repeat two sentences as accurately as possible. The first sentence, *The man saw the body that the dog chased* (“Man Dog”), was read with an overall accuracy of 76.15% (SD = 30.28%). In response to this passage, participants aged 18-39 (M = 77.75%, SD = 29.34%) read passages significantly more accurately than did participants aged 40-69 (M = 71.55%, SD = 32.42%), *t*(2459) = 4.452, Bonferroni-corrected *p* = 0.00016. There was no evidence of difference between genders in this task.

In response to the second sentence, *The tour bus is coming into the town to pick up the people from the hotel to go swimming* (“Tour Bus”), participants performed with an average accuracy of 69.63% (SD = 30.00%). Once again, participants aged 18-39 (M = 70.70%, SD = 29.40%) read passages significantly more accurately than participants aged 40-69 (M = 66.57%, SD = 31.49%), *t*(2458) = 2.989, Bonferroni-corrected *p* = 0.051. There was no evidence of difference between genders in response to this second sentence.

### Speech rate

The various speech tasks in the Voiceome Dataset resulted in different speech rates, as measured by the number of voice segments per second from the OpenSMILE GeMAPS embeddings. Figure 2 plots the average speech rate across each speech task. The speech rates are consistent with the idea that tasks which require substantial effort and cognitive load—such as the letter F naming task—result in comparatively lower speech rates, whereas less cognitively demanding tasks—like the Caterpillar passage task—result in comparatively higher speech rates. Another notable finding is that some of the immediate recall tasks differed in speech rate, which could be due to the length of the immediate recall task (e.g., the Man Dog task had 9 words and was 4 seconds long during playback, whereas the Tour Bus task had 14 words and was 7 seconds long during playback). In sum, the results denoted in Figure 2 indicate that speech rate appears to be a powerful feature to represent the relative cognitive load of speech-based survey tasks.

**Figure 2.**
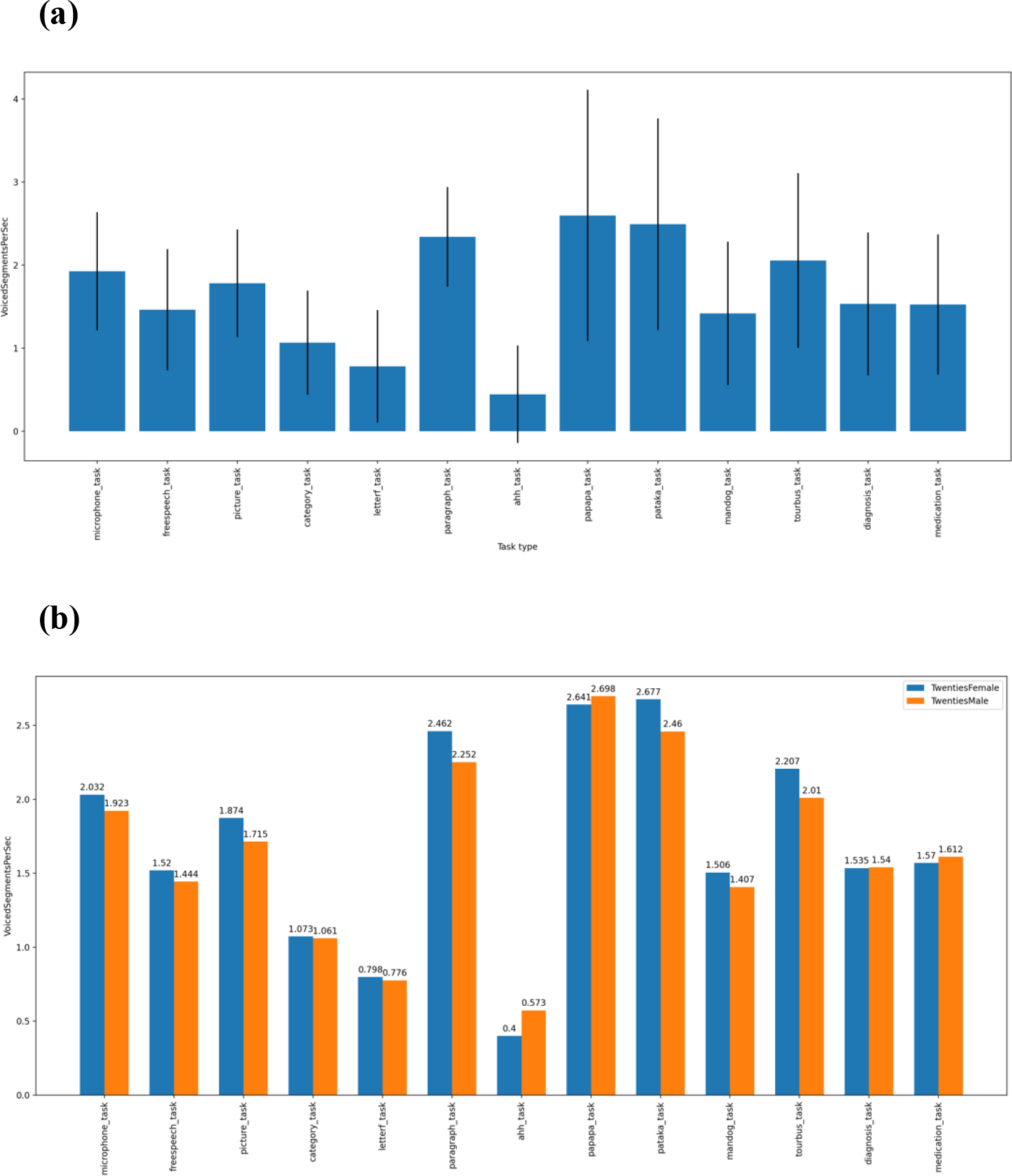
Participant speech rates across voice tasks for Survey A for (a) all participants and (b) comparing males and females aged 20-29. Both subplots represent the speech rate as represented by the VoiceSegmentsPerSec feature extracted from the OpenSMILE GeMAPS embedding. The speech tasks are represented in the order that participants completed the tasks in the Voiceome Dataset. For example, the ‘00_mic_check’ label corresponds with the microphone task, which was the first voiced question subjects were asked to complete, and the 49_medication_task label corresponds with the spoken medication task, which was the last voiced questions subjects were asked to complete. Customized graphs of speech rates by gender and age cohorts—such as subplot (b)—can be created using the Voiceome GitHub (https://github.com/jim-schwoebel/voiceome).

#### Free speech

In response to the free speech task, *Tell us about a recent happy memory based on experiences from the past month*, participants spoke with an average speech rate of 90.20 words per minute (SD = 39.86). There was no evidence of difference in overall speech rate between females and males or between individuals 18-39 years of age and 40-69 years of age.

Brunet’s index (Brunet, 1978) and Honoré’s statistic (Honoré, 1979) are both metrics that quantify lexical richness used in speech. Lower values of Brunet’s index indicate more speech richness which are generally independent to text length (normal ranges is 10.0-20.0; Holmes & Singh, 1996), whereas higher values of Honoré’s statistic indicate more speech richness. Overall, Voiceome participants had an average of 9.81 for Brunet’s index (SD = 1.69) and 1875.42 for Honoré’s statistic (SD = 828.81). Once again, there was no evidence of difference in Brunet’s index or Honoré’s statistic between females and males or between people aged 18-39 and people aged 40-69.

Figure 3 further explores speech characteristics in response to the free speech prompt. In this graph, the visible gap between the fundamental frequency distribution of self-reported males and females appears to get smaller as people grow older. Speech rate, defined by number of voiced segments per second, may be seen to decrease as people age; similarly, the average duration of pauses when speaking may tend to increase over the course of one’s lifetime.

**Figure 3.**
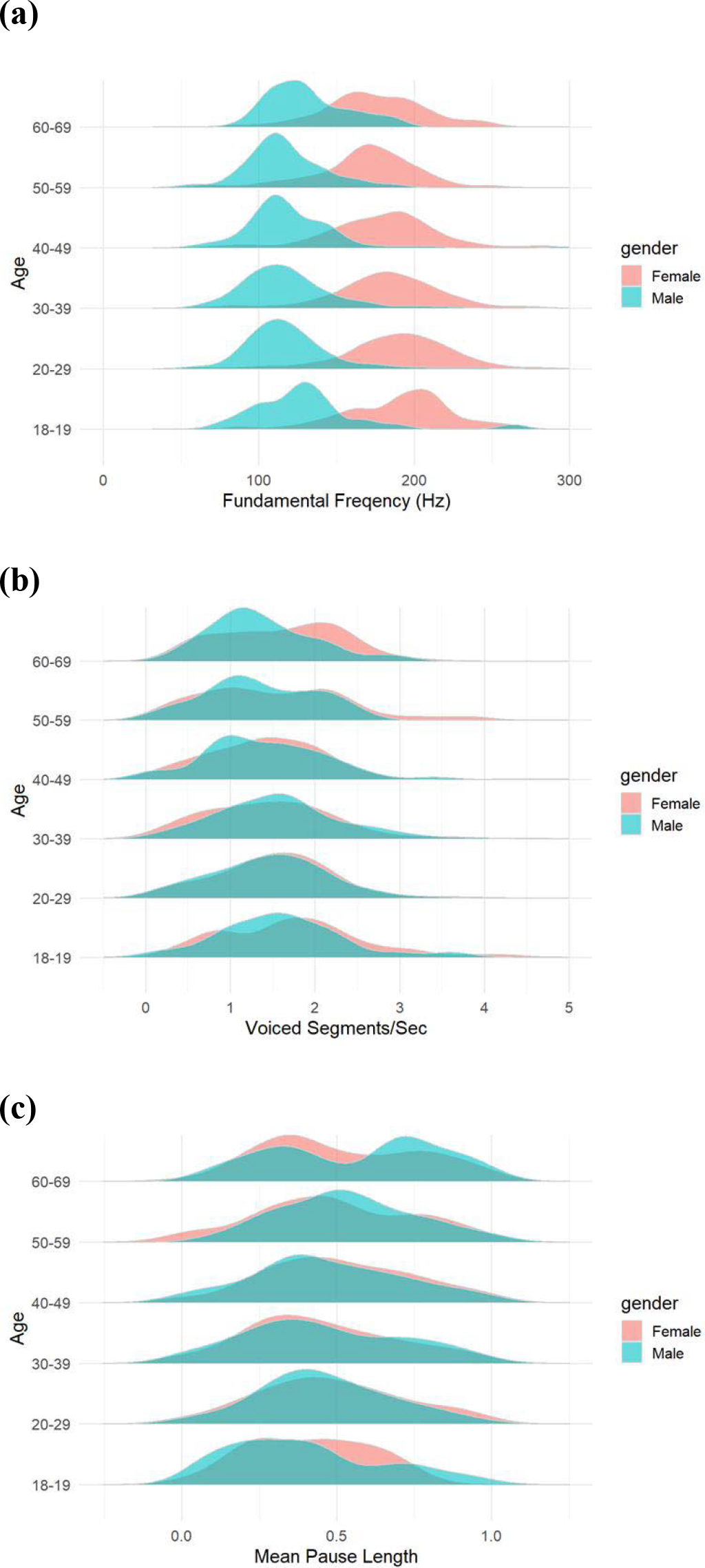
Distributions of speech features of Voiceome participants in response to the free speech prompt, “Tell us about a recent happy memory based on experiences from the past month” by gender and age (in decades). (a) fundamental frequency, (b) speech rate, (c) average pause length.

#### Picture description

Participants were shown an image (reproduced in Table 4) and asked to, *Tell us everything you see going on in this picture*. Overall, participants responded with an average speech rate of 115.34 words per minute (SD = 37.87). Females (M = 116.981, SD = 36.79) spoke slightly faster than males (M = 112.08, SD = 39.61), on average, *t*(2377) = 3.012, Bonferroni-corrected *p* = 0.047. There was no evidence of difference in speech rate between people 18-39 and 40-69 years old.

Voiceome participants had an overall average of 10.82 for Brunet’s index (SD = 1.65) and 1696.01 for Honoré’s statistic (SD = 475.75). Males spoke with comparatively more lexical richness than females, as measured by Honoré’s statistic (females: M = 1657.74, SD = 421.33; males: M = 1769.98, SD = 558.25; *t*(2377) = -5.506, Bonferroni-corrected *p* < 0.001), but this difference was not evident with Brunet’s index. There was no evidence of difference in lexical richness between participants ages 18-39 and 40-69 using Honoré’s statistic or using Brunet’s index.

#### Phonetically-balanced paragraph reading

Voiceome Dataset participants were asked to read the Caterpillar passage (Patel et al., 2013). Due to a technical error, participants’ speech recording ended at 60 seconds, whereas this passage generally takes about 90 seconds to read. During the first minute of speech, participants spoke at a rate of 162.25 words per minute (SD = 34.19). Participants between ages 40-69 read the passage significantly slower (M = 157.70, SD = 32.21) than did participants between ages 18-39 (M = 163.83, SD = 34.71), *t*(2380) = 3.833, Bonferroni-corrected *p* = 0.002. There was no evidence of difference between the speech rate of females and males.

#### Repeating syllables

When asked to repeat the syllables *pa-pa-pa*, participants spoke at an average rate of 2.60 voice segments per second (SD = 1.52). There was no evidence of difference in the rate of speech between females and males or between individuals 18-39 and 40-69 years old.

When asked to repeat the syllables *pa-ta-ka*, participants spoke at an average rate of 2.49 voice segments per second (SD = 1.28). There was no evidence of difference in the rate of speech between females and males or between participants aged 18-39 and 40-69.

#### Non-word task

Participants were asked to pronounce 10 “non-words” (e.g., *plive*, *fwov*, *zowl*), a speech task unused in SLB digital research, but previously examined as a test to dissociate mechanisms of reading in Alzheimer’s disease (Brain and Language, 43, 400-413, 19912, Friedman, Ferguson. Robinson). Of these ten words, participants correctly pronounced an average of 5.33 (SD = 0.62) words. On average, this task took an average of 30.28 seconds (SD = 26.43) to complete. There was no evidence of difference in duration by gender or by age.

### Naming tasks

#### Category naming

Participants were asked to produce as many animals as they could think of within 60 seconds. On average, participants named 18.85 animals (SD = 9.89) within one minute. There was no evidence of difference between females and males or between participants ages 18-39 and 40-69.

#### Phonemic fluency

In order to measure phonemic fluency, participants were asked to produce all the words beginning with the letter F that they could think of within 60 seconds. Overall, participants named an average of 14.82 F words (SD = 6.16). Females (M = 15.14, SD = 6.16) named slightly more F words than did males (M = 14.17, SD = 6.40), *t*(2417) = 3.659, Bonferroni-corrected *p* = 0.005. There was no evidence of difference between participants 18-39 years old and 40-69 years old.

#### Confrontational naming

Participants were shown images of objects (e.g., mushroom, bicycle) and were asked to speak the name of the object within 10 seconds. In total, there were 25 images. On average, participants named an average of 17 (SD = 3.53) of the 25 words correctly.

To complete the entire confrontational naming section, participants had average total duration of 76.78 seconds (SD = 61.27). Males (M = 82.00, SD = 65.20) were faster at this task than females (M = 74.01, SD = 58.84), *t*(2461) = -3.091, Bonferroni-corrected *p* = 0.036. There was no evidence of difference across age groups in average duration.

### Sustained phonation

Participants were asked to make the vowel sound “/a/” (as in hallelujah) for as long as they could during a 30-second timer. The average phonation time for all participants was 19.43 (SD = 7.16) seconds in duration. Females (M = 18.51, SD = 7.01), on average, had shorter sustained phonation times than did males (M = 21.11, SD = 7.11), *t*(1850) = -7.61, Bonferroni-corrected *p* < 0.001. There was no evidence of difference in average phonation time between individuals aged 18-39 years and 40-69 years. Table 5 delineates differences in sustained phonation by age (in decades) and gender.

**Table 5.** Duration of sustained phonation to the vowel ‘/a/’ by gender and age.

| *Gender* | *Age* | *n* | *Mean* | *SD* |
| --- | --- | --- | --- | --- |
| Female | 13-19 | 45 | 18.272 | 6.953 |
|  | 20-29 | 444 | 18.801 | 6.774 |
|  | 30-39 | 364 | 18.851 | 7.160 |
|  | 40-49 | 171 | 17.286 | 6.828 |
|  | 50-59 | 100 | 17.817 | 7.435 |
|  | 60-69 | 61 | 19.088 | 7.446 |
| Male | 13-19 | 19 | 19.090 | 6.540 |
|  | 20-29 | 269 | 20.960 | 7.100 |
|  | 30-39 | 230 | 21.543 | 7.176 |
|  | 40-49 | 88 | 21.652 | 6.764 |
|  | 50-59 | 34 | 20.959 | 7.830 |
|  | 60-69 | 27 | 18.637 | 6.970 |

### Comparing the novel non-word speech task with the Boston Naming Test task

Both the non-word speech task and confrontational naming task (used in the Boston Naming Test; Kaplan, Goodglass, & Weintraub, 1983) had similar instructions. Either a text string—for the non-word task—or a picture—for the confrontational naming task—were presented on the screen; participants were asked to speak single word responses to what they saw on the screen (see the Methods section for more details on these task instructions). Participant responses to these two tasks were compared in several ways (Figures 4-6 below).

**Figure 4.**
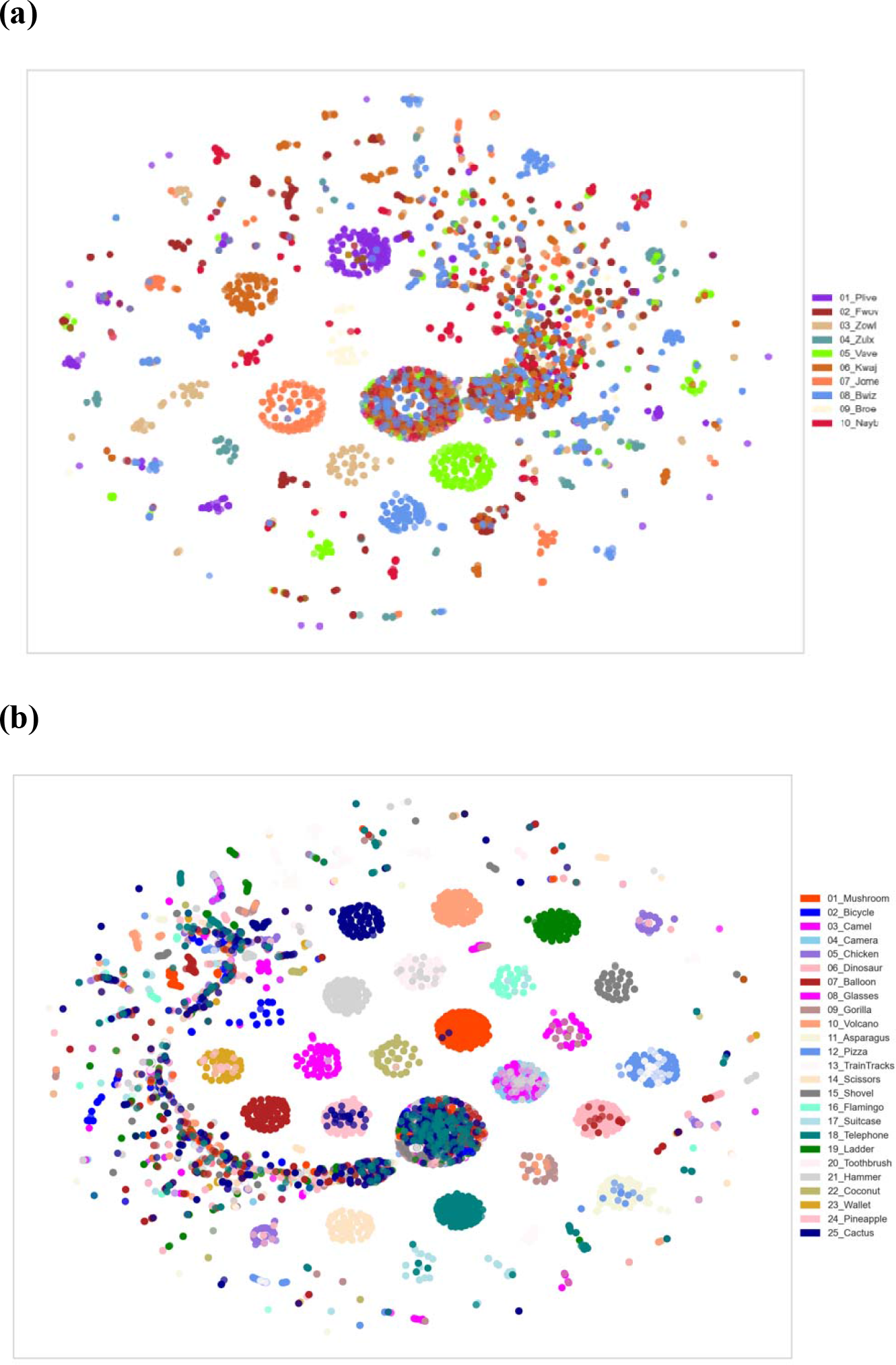
t-SNE plots from Survey A responses for the (a) non-word naming task and (b) confrontational naming task. The t-SNE plot for the non-word naming tasks shows that, despite multiple variants of elicitations for each non-word (e.g., ‘bwiz’ has 2-3 clusters), some non-words are distinct from other non-words, whereas other non-words overlap with other non-words. The t-SNE plot for all confrontational naming tasks contains >20 clusters, demonstrating the independence of each picture task and indicating high task compliance. Most overlap between these tasks is due to the use of determiners (e.g., “the” dinosaur, “a” shovel). All t-SNE plots represented used Azure transcripts as the source reference and were generated with the t-SNE Corpus Visualization feature in Yellowbrick: https://www.scikit-yb.org/en/latest/api/text/tsne.html

Figure 4 shows t-SNE plots for these two tasks (confrontational and non-word naming). Recall that t-SNE can be used to uncover the number of independent clusters from a series of speech tasks or prompts; t-SNE therefore provides a visual metric regarding the similarity or difference among participant responses to the speech tasks. For the confrontational naming task, which contained 25 images, the t-SNE plot reveals more than 20 clusters. The corresponding interpretation is consistent with the idea that each picture task resulted in independent clusters, therefore indicating that participants highly complied with the task instructions. Post-hoc analysis shows that the use of speech determiners like *a* or *the* sometimes resulted in some cluster overlap. The t-SNE plot for the non-word naming tasks is also consistent with the idea that there is dimensional independence for each non-word. In some cases, the different pronunciations of non-words resulted in more than one cluster per non-word (e.g., *bwiz*). In general, though, each non-word resulted in a single cluster, even taking into account the multiple pronunciations by Voiceome participants.

Two measures of word complexity were used to compare participants’ performance on the confrontational naming task and the non-word task (Figure 5). Word complexity was first operationalized by looking at the five unique phrases that were spoken most frequently to describe the non-word text or the image (left side of Figure 5). The distribution of the frequency among the top five utterances indicates how similar participants pronounced the words. When the distribution is highly skewed towards the top word, such as the responses to non-word *broe*, it means that almost all participants pronounced the (non-)word the same way. When the distribution is more uniform across the five words, such as the responses to non-word *fwov*, participants used a variety of pronunciations to speak the (non-)word. In this case, *fwov* would be considered to be more complex than *broe*. As expected, the non-words were generally interpreted as more complex than the confrontational images.

**Figure 5.**
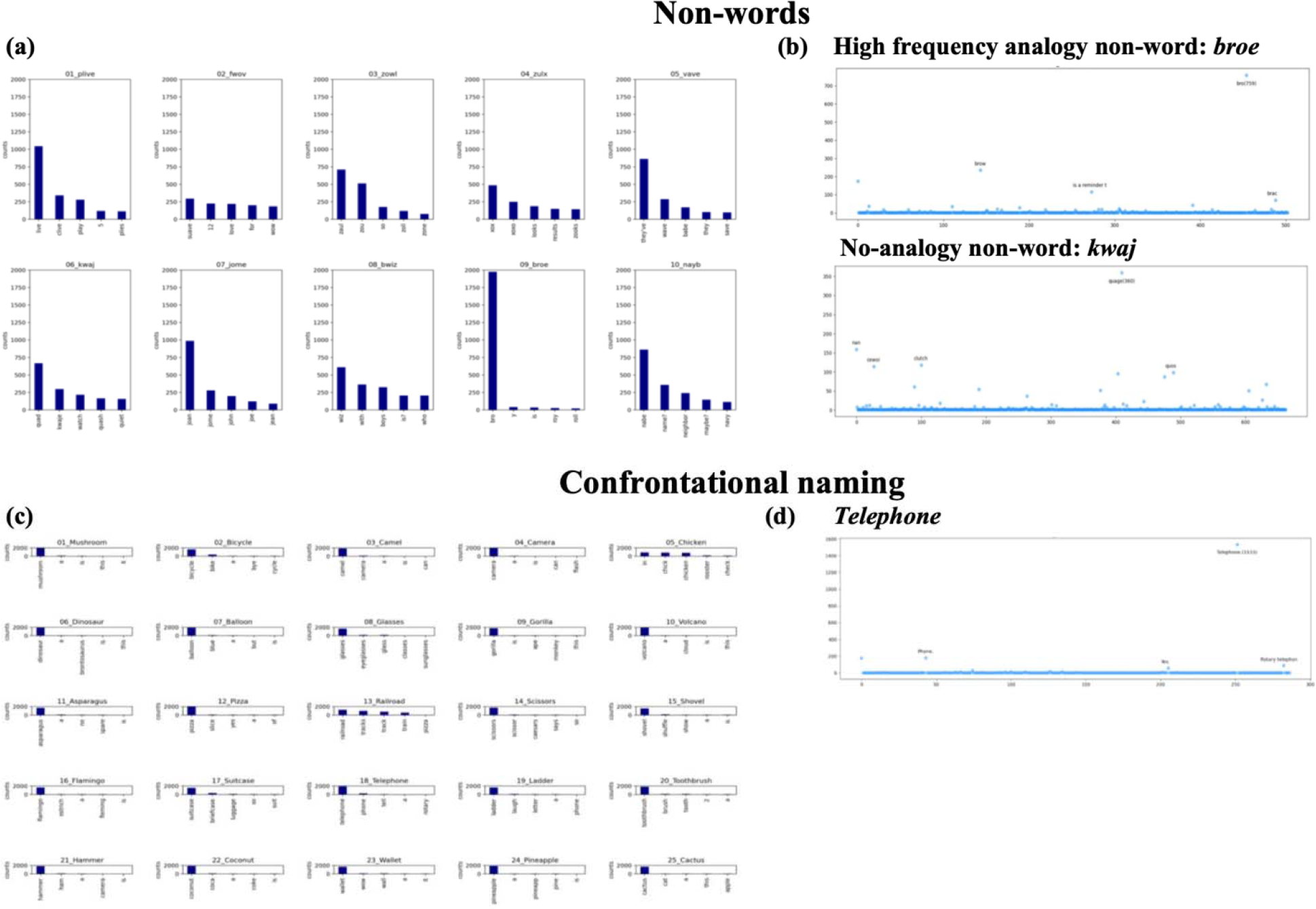
Two types of word complexity metrics for the non-word task (subplots A and B) and the confrontational naming task (subplots C and D). The diagrams on the left (subplots A and C) represent the number of times the top 5 unique phrases were used to describe words from the (a) non-word task and (c) confrontational naming task. The diagrams on the right (subplots B and D) provide examples of complexity for individual (non-)words, where the x-axis represents the number of unique utterances per word and the y-axis indicates the number of times each unique utterance was spoken by participants. For the word *telephone*, there were about 290 unique utterances, some of which included *telephone* (1,533 utterances), *rotary telephone* (about 100 utterances) and *phone* (about 200 utterances). Responses to the confrontational naming tasks, such as subplot D, were transcribed with Azure because Azure is based on English words. Responses to non-words, such as subplot B, were transcribed with HuBERT because HuBERT transcriptions are not based on any languages.

Word complexity was also defined as the total number of unique phrases spoken per non-word or image. As the number of descriptors per non-word/image increases, it may indicate that the non-word or image was harder to identify. For example, the non-word *zulx* has over 1,000 unique phrases whereas *jome* has roughly 500 unique phrases, making *zulx* roughly two times more complex of a non-word than *jome*. The right side of Figure 5 shows the total number of unique phrases participants spoke in response to the non-words *broe* and *kwaj* and to an image of a *telephone*. Here too, the non-words seem to be more complex than the confrontational images. An important caveat relates to the two types of non-words used in the study. Non-words with high frequency analogy to English words, such as *broe*, were less complex than non-words with no analogies to English words, such as *kwaj*. Additional information about extracted acoustic features from this task is provided in Table C.1 in the Supplemental Materials.

Figure 6 shows the energy of a person’s voice over the course of their recorded speech. One finding from this plot is that participants took longer to start speaking after viewing the Boston Naming Test images than when they saw the non-words on the screen. One possible interpretation of this finding could be that there is a higher cognitive load for the Boston Naming Test because it involves cross-modal associations between multiple modalities (e.g., visuospatial input, memory retrieval, and speech articulation), while the non-word task results in a lower cognitive load because it mostly involves strategies of text reading (reading text and speaking words).

**Figure 6.**
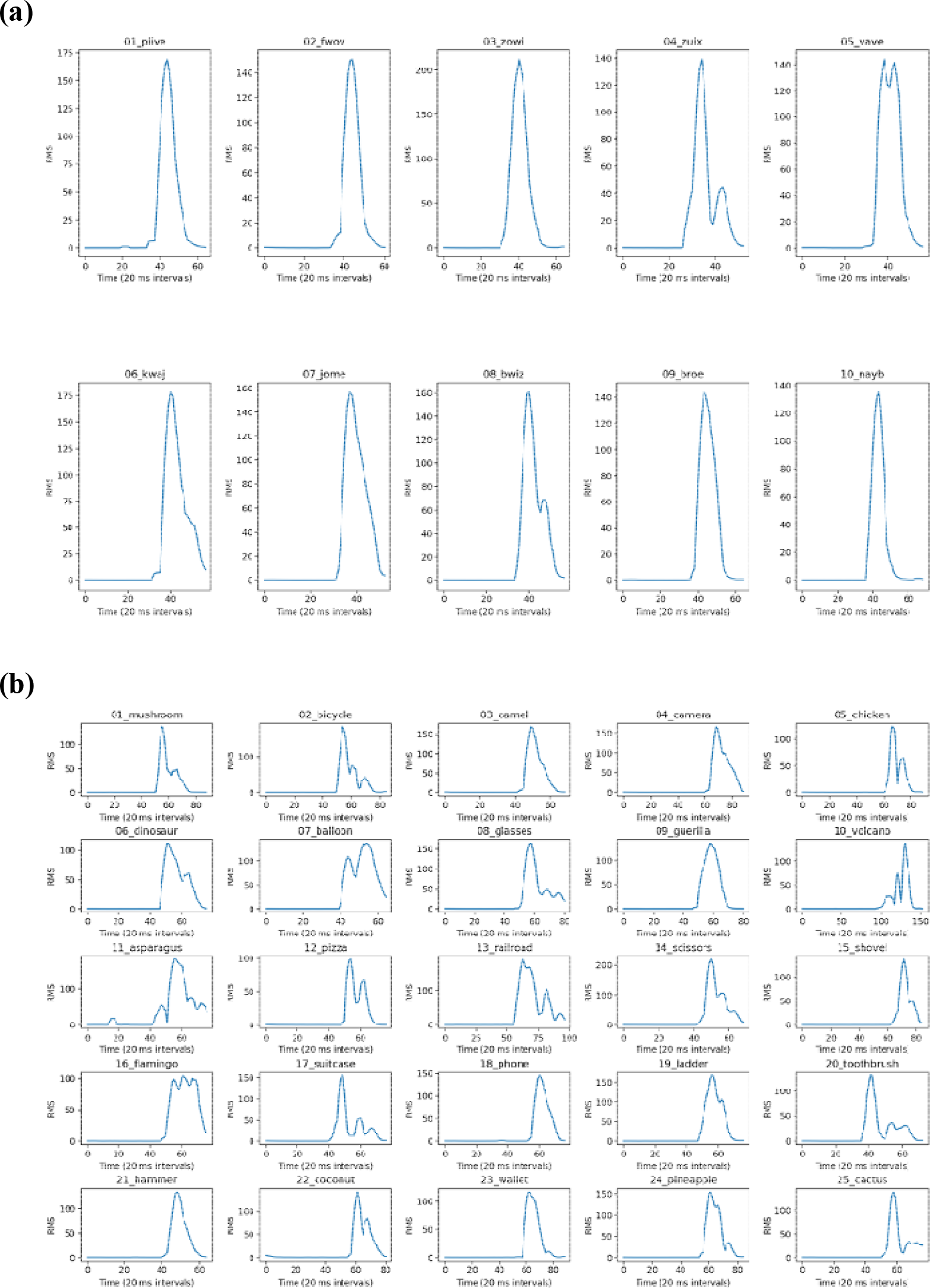
Amount of energy in speech over time for (a) non-words and (b) confrontational naming images. Energy is measured as root mean square (RMS) values.

**Figure 7.**
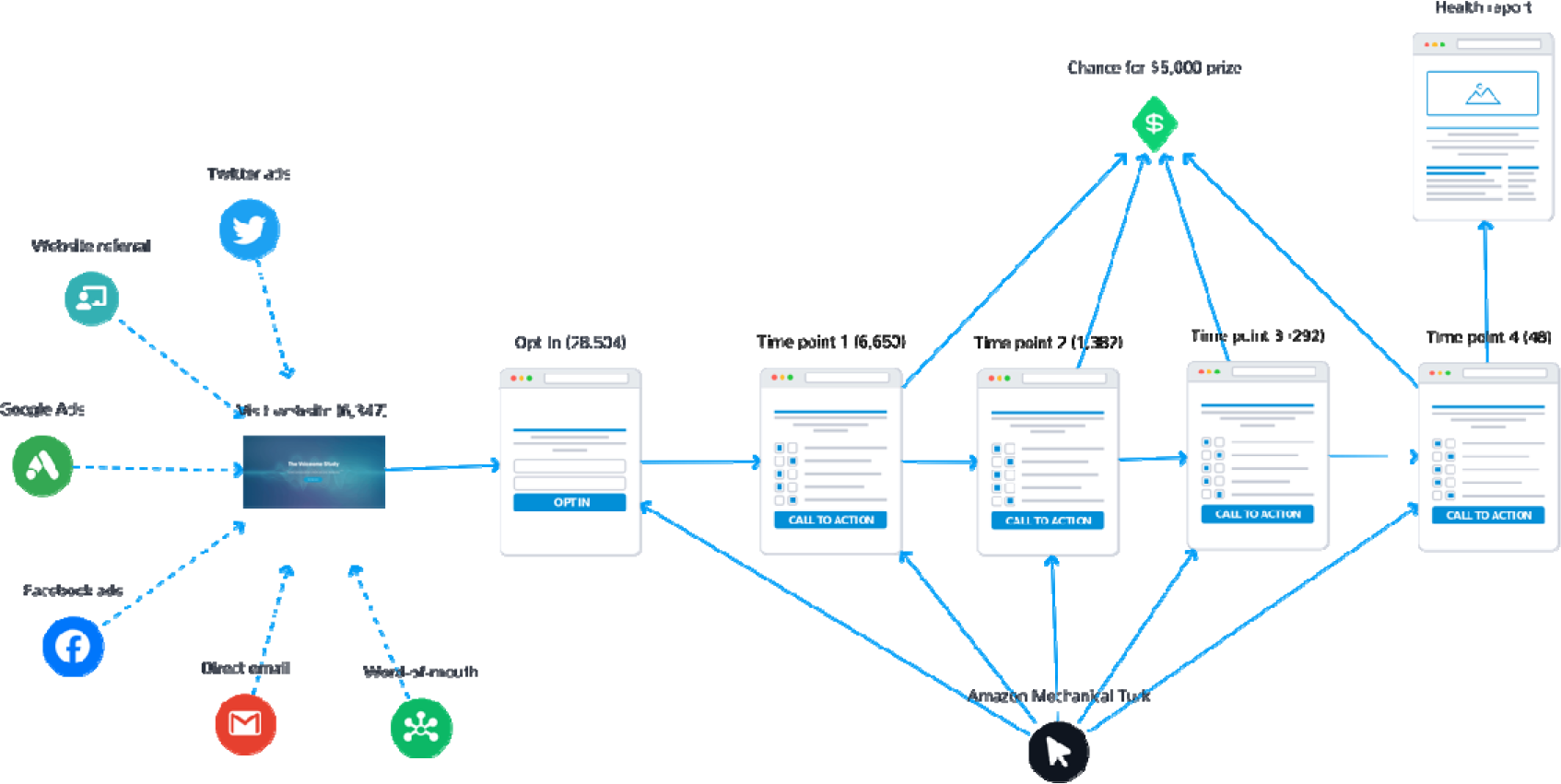
Voiceome Dataset recruitment methods. This figure shows the typical funnel for a Voiceome Dataset participant. First, users visit the Voiceome.org website and opt-in to the clinical study via a survey form. Then users fill out the first survey and are reminded via text messages and email reminders to follow up in the following weeks. As shown above, many recruitment methods were used including direct email outreach, word-of-mouth referral, Amazon Mechanical Turk (mTurk), Facebook ads, Google ads, website referrals (from embeddings), and Twitter ads; however, the most effective method for recruiting clearly was mTurk with a $5-10 incentive for each survey completed. Overall, over 28,000 participants opted in to the study, 6,650 participants completed the first time point, 1,382 participants completed the second time point, 292 participants completed the third time point, and 48 completed the last time point. Overall, this proves that mTurk can be reliably used as a source of recruiting for decentralized trials related to SLBs, resulting in high-quality and quick completion of trials.

## Discussion

The Voiceome Study covers two aspects of SLB research. First, the Voiceome Protocol offers a scalable speech and language biomarker (SLB) protocol with twelve kinds of neuropsychological speech and language assessments that researchers and clinicians can easily use to conduct large-scale and decentralized research. Second, the corresponding Voiceome Dataset provides normative SLB performance metrics for over six thousand participants. These participants are broadly representative of the United States population. In total, there are five notable aspects of the Voiceome Study, all of which are detailed below.

### 1: Helps make SLB research accessible to both researchers and participants

As demonstrated above, the Voiceome Study offers a scalable and accessible protocol that can be easily used by other researchers. The protocol can be applied widely across many health conditions, including neurological and motor coordination conditions (e.g., Alzheimer’s disease, Parkinson’s disease, stroke, intoxication), mental health conditions (depression, schizophrenia, anxiety), and respiratory conditions (asthma, COPD, COVID-19). Given the fact that the speech performance metrics presented in this paper are broadly representative of the United States population, researchers can use the Voiceome Dataset benchmarks as metrics with which to compare clinical populations.

Researchers can use the SurveyLex platform (https://www.surveylex.com) to duplicate (and modify) the Voiceome Protocol in less than one minute. The Voiceome templates on SurveyLex include all twelve speech tasks, as well as all demographic & health questions and questions relating to common speech-research confounds. Given that clinical tests often take a long time to administer (>1-2 hours), require an expert to collect the data (e.g., a neurologist or a nurse), and require in-person measurements (e.g., clinic, hospital), the Voiceome Protocol offers a reliable and reproducible source of data at a significant cost and time savings relative to these clinical alternatives. By distributing the Voiceome Protocol online instead of in person, study completion time can drop from 2 hours to 20 minutes, a time savings of up to sixfold. Likewise, compared to the cost of an in-person study, roughly 200 USD per participant, the same study deployed on SurveyLex would only cost 20 USD per participant, a tenfold savings in cost. The Voiceome Study furthermore demonstrates the utility of using SurveyLex for decentralized clinical studies even during unexpected global events such as the COVID-19 pandemic crisis.

SurveyLex allows researchers to download all study results, including survey responses and the speech recordings. In addition, the Voiceome GitHub (https://github.com/jim-schwoebel/voiceome) can be used to analyze the data from any study using the Voiceome Protocol. The GitHub allows researchers to listen to their participants’ recordings. For each recording, the GitHub can be used to create feature embeddings for spectral and prosodic acoustic features, pause detection, and text analysis of the transcript. Furthermore, the GitHub can help provide numerical performance metrics for each of the twelve speech tasks presented in this paper, as well as the performance benchmarks for the Voiceome Dataset participants. As mentioned above, the GitHub can be used to compare SLB benchmarks for different participant cohorts, such as “males from ages 20-29.” Through this feature, researchers can compare their patient population with a matched Voiceome Dataset cohort.

The Voiceome Protocol demonstrates the feasibility of collecting health data online, allowing researchers to reach larger populations, connect with people suffering from a disease from the comfort of their own homes, and to easily collect data for underrepresented individuals in the clinical literature (e.g., bilingual speakers). The implications of applying the Voiceome

Protocol to clinical populations affect many aspects of healthcare. The protocol can be used in a cross-sectional manner to compare various patient populations to the Voiceome Dataset benchmarks. The Voiceome GitHub allows researchers to easily match speech metrics for their clinical population with normative benchmarks with regard to age, gender, language, accent, and more. The Voiceome Protocol can furthermore be used for tracking a patient’s health over time. Early symptom detection and symptom monitoring over time is made possible by measuring an individual’s speech and language biomarkers in a longitudinal manner. In conclusion, the Voiceome Study facilitates both preventative and active health treatments, as well as the investigation of a plethora of health conditions for which speech and language biomarker research is novel.

### 2: Utilizes novel speech tasks and evaluation metrics

In addition to pioneering a method to collect SLB data digitally, the Voiceome Study offers novel forms of digitalized speech tasks and performance metrics. The Voiceome Protocol is the first survey to digitally utilize the *non-word speech task* in speech and language digital biomarker-related research. Previously and in analogue mode, the importance of spelling-to-sound correspondence was investigated by Friedman and colleagues, as a reading test to discriminate individuals with Alzheimer’s from normal controls (Friedman, Ferguson, Robinson, & Sunderland, 1992). In that study, individuals with Alzheimer’s disease (AD) were markedly impaired relative to the healthy controls in reading pseudowords with no analogues.

In the Voiceome Study non-words task, participants saw a series of pseudowords appear on the screen and were recorded as they pronounced the words out loud. Some of these non-words were designed to be similar to words in the English language (high frequency analogy non-words: *plive*, *zowl*, *vave*, *jome*, *broe*), whereas other non-words had no similar English neighbors (no-analogy non-words: *fwov*, *zulx*, *kwaj*, *bwiz*, *nayb*). As demonstrated in Figures 4-6, the results for the Voiceome Dataset are consistent with the idea that the high frequency analogy non-words had less variability in pronunciation than did the no-analogy non-words (Figure 5).

This clear separation can be useful in classifying individuals with Alzheimer’s disease vs healthy controls, as it has previously reported (Friedman et al., 1992), and this remains to be confirmed with test data. Even so, the results are consistent with the idea that there is dimensional independence for each non-word, regardless of its analogy with English words (Figure 4). Although further analytics are necessary for establishing better evidence, these results can be used in future dementia classification studies. For example, by selecting pseudoword *broe* (high frequency analogy) and pseudoword *fwov* (no analogy), these results provide indication that in an AD versus healthy controls classification test, an individual with AD is expected to answer with the most common response when tested for *broe*, and the same individual is expected to fail in reproducing any of the five natural distribution responses

Comparison between the non-word task and the confrontational naming task suggests that the non-word task is a robust alternative to the Boston Naming Test (BNT; Kaplan, Goodglass, & Weintraub, 1983), particularly for the dementia disease area, where individuals with Alzheimer’s perform rather poorly on the group of low frequency non words (Friedman, Ferguson, Robinson, & Sutherland, 1992). The non-word task may have benefits that extend beyond the BNT, as the non-word task may be easily adapted to other languages and can be used with participants of various levels of English fluency. Open-source automated transcription packages, such as DeepSpeech (https://github.com/mozilla/DeepSpeech/releases/tag/v0.7.0) or the novel HuBERT method (Hsu, Bolte, Tsai, Lakhotia, Salakhutdinov, & Mohamed, 2021) can reliably assess a person’s pronunciation of non-words, further extending the benefits of the non-word task to future SLB researchers. Future work should continue to explore non-word speech tasks to create a longer list of non-words to use as keyword dictionaries in SLB tasks. Similarly, keyword spotting algorithms could perhaps be used for non-words to make detection more robust into the future.

### 3: Offers new health information, including common confounds

The Voiceome Study also offers a rich protocol to screen for confounding factors related to SLB-related research studies. Health-related factors that would otherwise be clinically unobserved, such as corrective vision, dental issues, smoking history, and hearing impairments, are known to impact speech and language research. For example, if a person cannot clearly read text on a screen, such as the Caterpillar task, their overall speech error rate may increase and their overall speech rate may decrease relative to their speech when wearing corrective lenses. Dental issues, exposure to radiation, and having a chronic history of smoking may alter speech production through changes with regard to precision of articulation and timbral or spectral changes of their voice. By including self-reported data, we acquire a personalized health profile and weigh factors that can influence critical metrics.

It has also been shown that highly educated individuals produce higher type token ratios (Hübner et al., 2018) and larger number of unique words in tasks like verbal fluency (Kawano, Umegaki, Suzuki, Yamamoto, Mogi, & Iguchi, 2010), so it is important to control for factors such as socioeconomic status in any SLB data analysis. Some epidemiological studies have reported faster cognitive decline in more educated people (Teri, McCurry, Edland, Kukull, & Larson, 1995; Scarmeas, Albert, Manly, & Stern, 2006), whereas other studies report slower decline in individuals with Alzheimer’s disease who have attained more education (Fritsch, McClendon, Smyth, & Ogrocki, 2002).

By offering these types of questions in the Voiceome Protocol on SurveyLex (https://www.surveylex.com), other SLB researchers can control for these confounds in their research. By using the same wording and question format for these variables across studies, it increases the robustness of comparing results from new studies with the results from the Voiceome Dataset.

### 4: Illuminates new SLB findings

The Voiceome Dataset consists of responses from over six thousand participants who completed the surveys from the Voiceome Protocol. For each of the twelve speech tasks in the Voiceome Protocol, participants’ speech was analyzed according to standard SLB clinical guidelines. One important finding was that participants’ speech rates tended to vary among the different types of speech tasks. The difference in speech rates across task type was present when averaging the entire participant sample (Figure 2.A), as well as when examining different participant cohorts, such as males and females in their twenties (Figure 2.B).

Tasks such as semantic and phonemic fluency, which are known to activate memory retrieval, executive control, and other attention functions, result in comparatively lower speech rates than in tasks that require lower cognitive load, such as the caterpillar passage or the diadochokinesis tasks (*pa-pa-pa*, *pa-ta-ka*), where the speech rate is comparatively higher. Future research should directly evaluate the cognitive load among the twelve speech tasks presented here, especially when conducting individualized and longitudinal follow ups with each patient. One possible result of this future study is that speech rate may be putatively a measure of cognitive load.

The results of the Voiceome Dataset are also consistent with the idea that participants had high task compliance across the twelve speech tasks present in the study. For example, speech samples from most images or words in the non-word task, the confrontational naming task, and the diadochokinesis tasks form distinct clusters in representative t-SNE plots, indicating that each picture or work elicited an independent speech response and that participants adhered to compliance.

### 5: Provides representative speech performance benchmarks

The Voiceome Dataset offers more than 300 analytic metrics for the twelve speech and language research tasks presented in the Voiceome Protocol. Many of the ranges and distributions for these SLB metrics were previously unknown in the research community or were unreported in research papers. Furthermore, the Voiceome Dataset results for all speech tasks that had been previously reported matched what was expected from the corresponding peer-reviewed normative clinical data. The replication of known speech metrics in the Voiceome Dataset suggests that the customized digital distribution platform (SurveyLex) and analysis software (Voiceome GitHub), as well as standardized automatic speech transcription methods (e.g., DeepSpeech, huBERT) and feature extraction software (e.g., Allie Repository, OpenSMILE) are promising avenues to conduct future SLB research, especially given that these tools may enable more affordable and accessible research for participants, clinicians, and researchers.

The Voiceome Dataset consists of speech responses and corresponding health and demographic information for 6,650 participants. The overall participant body was broadly representative of United States population, including variables such as age, gender, socioeconomic status, BMI, race and ethnicity, and prevalence of clinical depression and anxiety. In addition to containing the representative U.S. sample, the Voiceome GitHub (https://github.com/jim-schwoebel/voiceome) allows researchers to explore numeric and visual representations of speech metrics by defining a cohort of interest, including variables such as age, gender, location, and health condition. The GitHub also offers a description of each speech task, sample audio responses, and exact instructions used in the Voiceome Protocol surveys.

The results from the Voiceome Dataset can be used as normative standard benchmarks with which results from non-clinical populations can be compared. Any clinicians or SLB researchers studying clinical populations may also wish to compare patient populations with the Voiceome Dataset benchmarks, as the comparison may elucidate speech discrepancies among the clinical and non-clinical samples. Given that speech and language biomarkers can be indicative of a number of health conditions, including respiratory, neurological, motor incoordination, mental health, and intoxication, the breadth of the Voiceome Dataset’s potential scope seems wide. Indeed, the Voiceome Protocol is currently being used in targeted clinical studies, such as dementia and depression, in order to compare the speech of representative non-clinical participants with speech from condition-specific cohorts of people.

#### Limitations

There are limitations to the Voiceome Protocol survey design. Participants self-identified their own medical diagnoses and symptoms, which may affect some of the ground-truth health labels. Although some participants noted their medications (which gave greater confidence on their diagnoses), the distribution of self-reported medication differed from what is expected in terms of medication prevalence, possibly suggesting participants’ hesitancy to acknowledge that they were taking medications. Yet even in clinical settings, clinicians diagnose their patients by asking patient-reported questions and inevitably factors of uncertainty should be considered by the physician. Physicians or researchers may also wish to consider a balanced recruitment strategy in the future, in order to optimize for longitudinal retention.

The Voiceome Dataset was conducted during the height of the COVID-19 pandemic (March 2019 through May 2020). The free speech task vocabulary was biased with COVID-19 related terms, so it is possible that the natural language embeddings may be skewed compared to non-pandemic times. Additional confounds like weather patterns and allergies were not thoroughly screened and could have affected SLB-related acoustic features.

## Conclusion

The Voiceome Protocol and Dataset offer a high-fidelity and normative dataset, as well as a scalable protocol that can be used to advance SLB research. The results of the study demonstrate that the online survey platform SurveyLex can be used as a tool to scale decentralized SLB-related research on a large-scale (n = 6,650 participants). The feasibility and scalability of using SurveyLex provides researchers and clinicians with the opportunity to standardize data collection efforts across academic centers and pharmaceutical partners. Through the methods presented here, it may be possible to reduce the time to take the survey protocol from 2 hours to 20 minutes (6x time savings) and survey costs from ∼$200/participant to ∼$20/participant (10x cost savings). It is our hope that the Voiceome Protocol and normative speech metric standards presented here can act as a template for future SLB-related research studies. You can clone the Voiceome Protocol in less than a minute at https://surveylex.com.

## Data Availability

The four versions of the Voiceome Protocol can be found at the following SurveyLex links below. Researchers can easily clone these surveys for their own use by selecting the "templates" feature during the SurveyLex survey design process.
Survey A - https://app.surveylex.com/surveys/8a32cbb0-cc8a-11eb-9ea3-938cc8b6d71e
Survey B - https://app.surveylex.com/surveys/061da3f0-a637-11eb-bcc9-eba67643f616
Survey C - https://app.surveylex.com/surveys/a66494c0-a824-11ea-88c1-ab37bac1e1d4
Survey D - https://app.surveylex.com/surveys/53737620-a637-11eb-bcc9-eba67643f616
To help with maximal replicability of this study design, all digital assets (audio, images, and text prompts) used for the trial were sourced either from open access Google searches, custom created by our research team, or acquired from other peer-reviewed articles. These are all available at https://github.com/jim-schwoebel/voiceome.
The complete data from the Voiceome Dataset can be accessed via a commercial license. If you would like access to this data, please contact the corresponding author.

## Acknowledgements

We would like to acknowledge Biogen for sponsoring this study and providing extensive help in recruiting participants and vetting the Voiceome protocol. We thank Shibeshih Belachew (MD, PhD) for assisting the review and approval process of the manuscript within Biogen. We would also like to acknowledge all the Voiceome Dataset participants who have helped to advance this form of research. We would also like to acknowledge Reza Hosseini Ghomi (M.D./MSE) for editing the study protocol on SurveyLex and drafting many of the early Institutional Review Board (IRB) documents for the protocol. We would also like to thank Drew Morris and Russell Ingram for helping to create the SurveyLex web product platform.

## Author Contributions

Jim Schwoebel, Eleftheria Pissadaki, Joel Schwartz, and Roland Brown conceptualized the study. Jim Schwoebel, Eleftheria Pissadaki, Joel Schwartz, Roland Brown, Monroe Butler, and Mark Moss built the Voiceome Protocol and clinical study design. Jim Schwoebel and Austin New built the SurveyLex web platform. Jim Schwoebel, Lindsay Warrenburg, and Roland Brown conducted the data analysis. Jim Schwoebel created the Voiceome GitHub page. Lindsay Warrenburg and Jim Schwoebel wrote the manuscript. All authors edited the manuscript.

## Methods

### Speech Tasks

Twelve separate speech task activities were used in the Voiceome Dataset. Across all twelve tasks, each participant spoke a total of 48 unique speech utterances. These tasks were selected because they provide non-overlapping information about a person’s health, as defined by previous literature (Tables 6 and 7). In addition, participants were asked to speak any clinical diagnoses and medications they were taking. All tasks proceeded in the same order, identified numerically in the text below. The Voiceome GitHub (https://github.com/jim-schwoebel/voiceome) provides all code used for audio pre-processing, feature extraction, and automatic transcription.

**Table 6.**
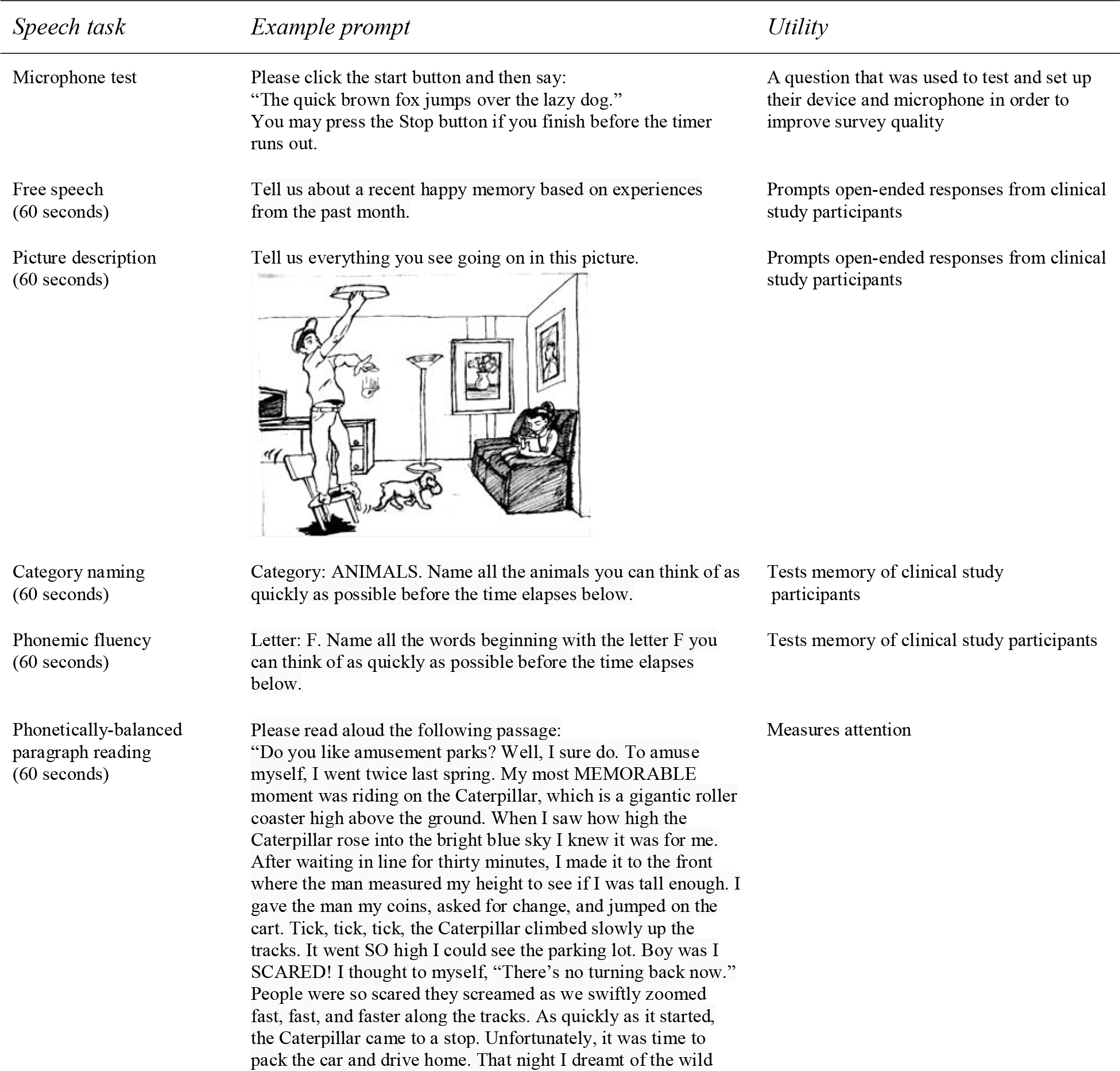
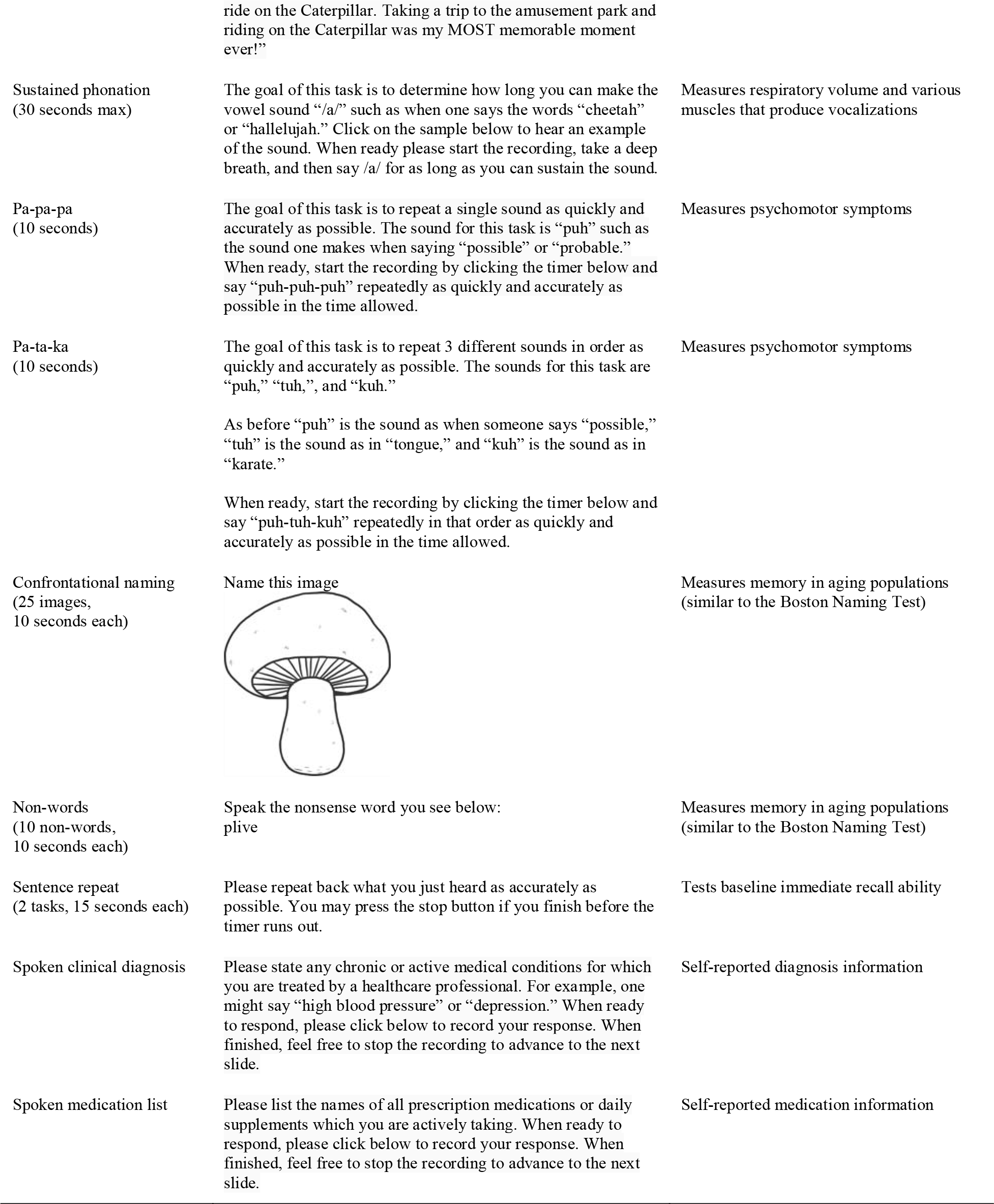
Speech tasks used in the Voiceome Dataset

| *Speech task* | *Example prompt* | *Utility* |
| --- | --- | --- |
| Microphone test | Please click the start button and then say:<br>“The quick brown fox jumps over the lazy dog.”<br>You may press the Stop button if you finish before the timer runs out. | A question that was used to test and set up their device and microphone in order to improve survey quality |
| Free speech<br>(60 seconds) | Tell us about a recent happy memory based on experiences from the past month. | Prompts open-ended responses from clinical study participants |
| Picture description<br>(60 seconds) | Tell us everything you see going on in this picture. | Prompts open-ended responses from clinical study participants |
| Category naming<br>(60 seconds) | Category: ANIMALS. Name all the animals you can think of as quickly as possible before the time elapses below. | Tests memory of clinical study participants |
| Phonemic fluency<br>(60 seconds) | Letter: F. Name all the words beginning with the letter F you can think of as quickly as possible before the time elapses below. | Tests memory of clinical study participants |
| Phonetically-balanced paragraph reading<br>(60 seconds) | Please read aloud the following passage:<br>“Do you like amusement parks? Well, I sure do. To amuse myself, I went twice last spring. My most MEMORABLE moment was riding on the Caterpillar, which is a gigantic roller coaster high above the ground. When I saw how high the Caterpillar rose into the bright blue sky I knew it was for me. After waiting in line for thirty minutes, I made it to the front where the man measured my height to see if I was tall enough. I gave the man my coins, asked for change, and jumped on the cart. Tick, tick, tick, the Caterpillar climbed slowly up the tracks. It went SO high I could see the parking lot. Boy was I SCARED! I thought to myself, “There’s no turning back now.” People were so scared they screamed as we swiftly zoomed fast, fast, and faster along the tracks. As quickly as it started, the Caterpillar came to a stop. Unfortunately, it was time to pack the car and drive home. That night I dreamt of the wild | Measures attention |
|  | ride on the Caterpillar. Taking a trip to the amusement park and riding on the Caterpillar was my MOST memorable moment ever!" |  |
| Sustained phonation<br>(30 seconds max) | The goal of this task is to determine how long you can make the vowel sound "/a/" such as when one says the words "cheetah" or "hallelujah." Click on the sample below to hear an example of the sound. When ready please start the recording, take a deep breath, and then say /a/ for as long as you can sustain the sound. | Measures respiratory volume and various muscles that produce vocalizations |
| Pa-pa-pa<br>(10 seconds) | The goal of this task is to repeat a single sound as quickly and accurately as possible. The sound for this task is "puh" such as the sound one makes when saying "possible" or "probable." When ready, start the recording by clicking the timer below and say "puh-puh-puh" repeatedly as quickly and accurately as possible in the time allowed. | Measures psychomotor symptoms |
| Pa-ta-ka<br>(10 seconds) | The goal of this task is to repeat 3 different sounds in order as quickly and accurately as possible. The sounds for this task are "puh," "tuh," and "kuh."<br>As before "puh" is the sound as when someone says "possible," "tuh" is the sound as in "tongue," and "kuh" is the sound as in "karate."<br>When ready, start the recording by clicking the timer below and say "puh-tuh-kuh" repeatedly in that order as quickly and accurately as possible in the time allowed. | Measures psychomotor symptoms |
| Confrontational naming<br>(25 images,<br>10 seconds each) | Name this image | Measures memory in aging populations (similar to the Boston Naming Test) |
| Non-words<br>(10 non-words,<br>10 seconds each) | Speak the nonsense word you see below:<br>plive | Measures memory in aging populations (similar to the Boston Naming Test) |
| Sentence repeat<br>(2 tasks, 15 seconds each) | Please repeat back what you just heard as accurately as possible. You may press the stop button if you finish before the timer runs out. | Tests baseline immediate recall ability |
| Spoken clinical diagnosis | Please state any chronic or active medical conditions for which you are treated by a healthcare professional. For example, one might say "high blood pressure" or "depression." When ready to respond, please click below to record your response. When finished, feel free to stop the recording to advance to the next slide. | Self-reported diagnosis information |
| Spoken medication list | Please list the names of all prescription medications or daily supplements which you are actively taking. When ready to respond, please click below to record your response. When finished, feel free to stop the recording to advance to the next slide. | Self-reported medication information |

**Table 7.**
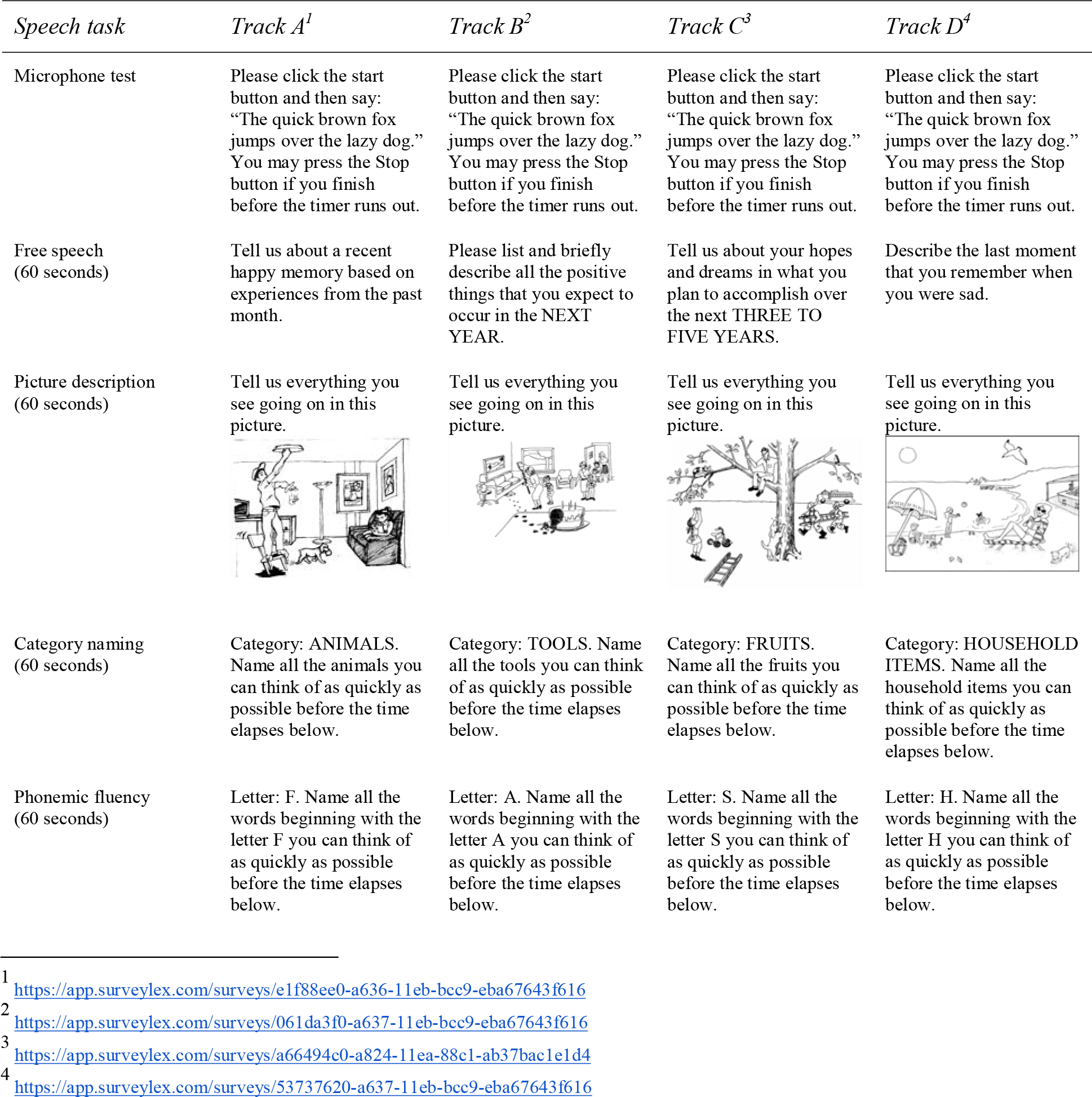
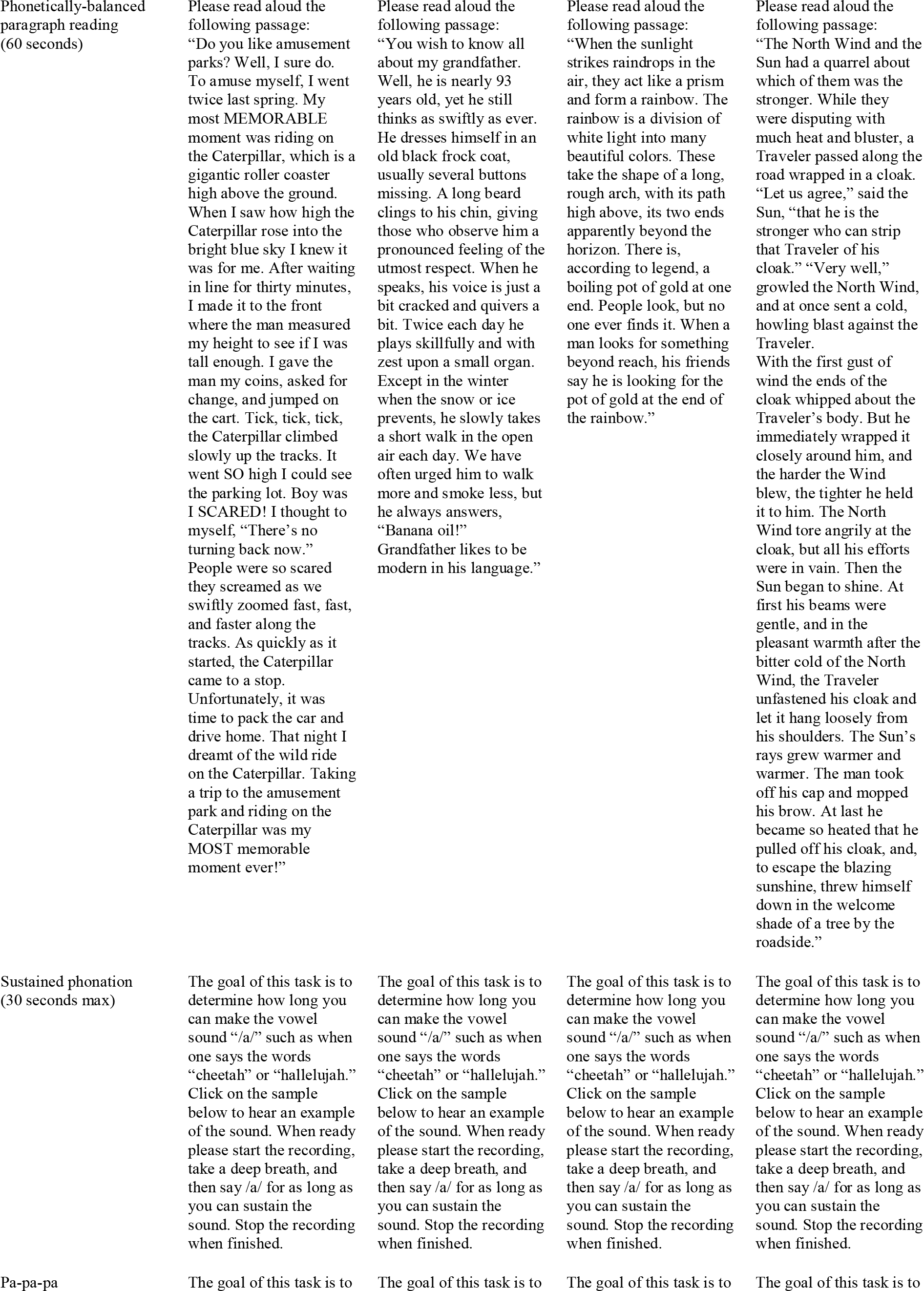
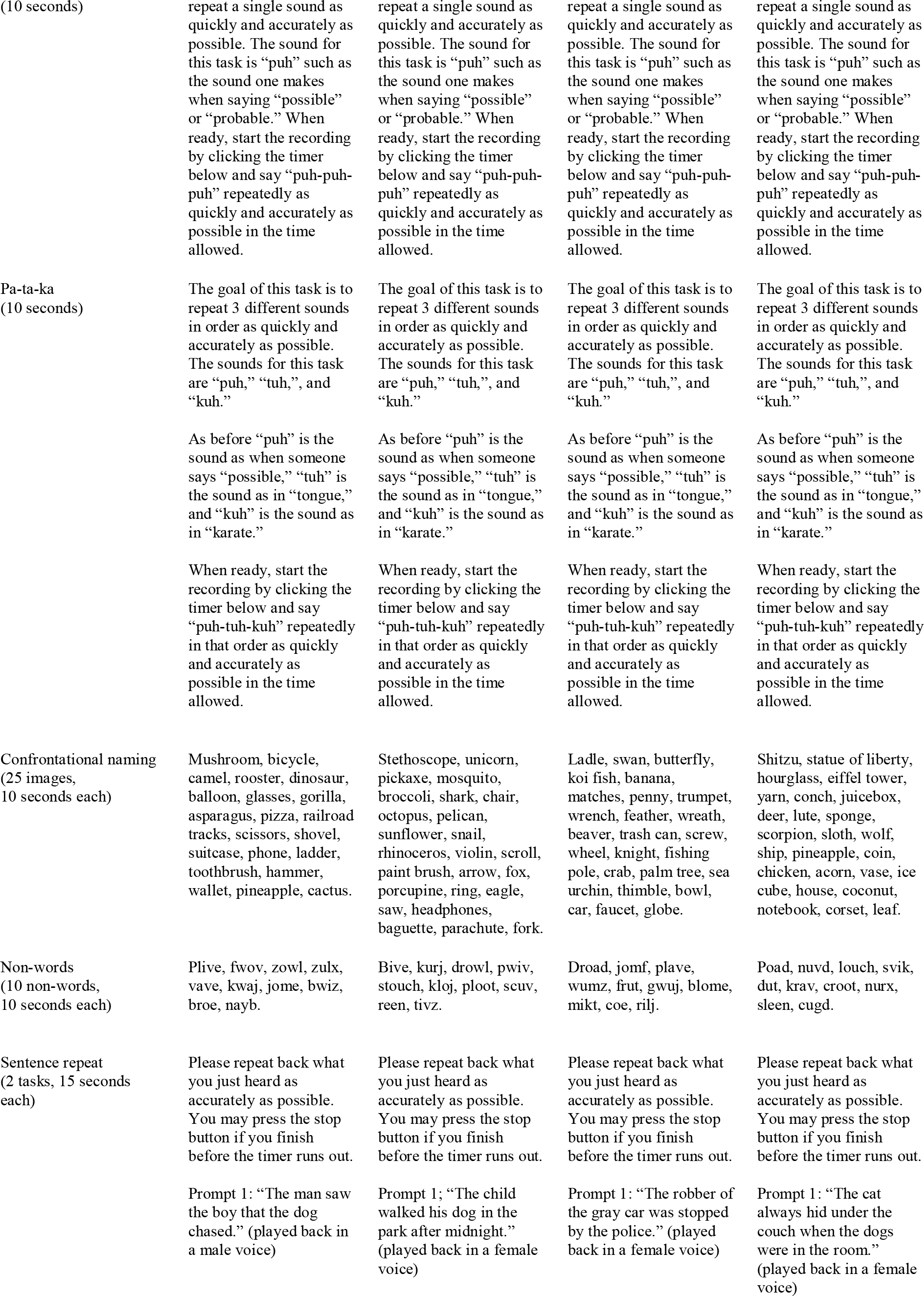
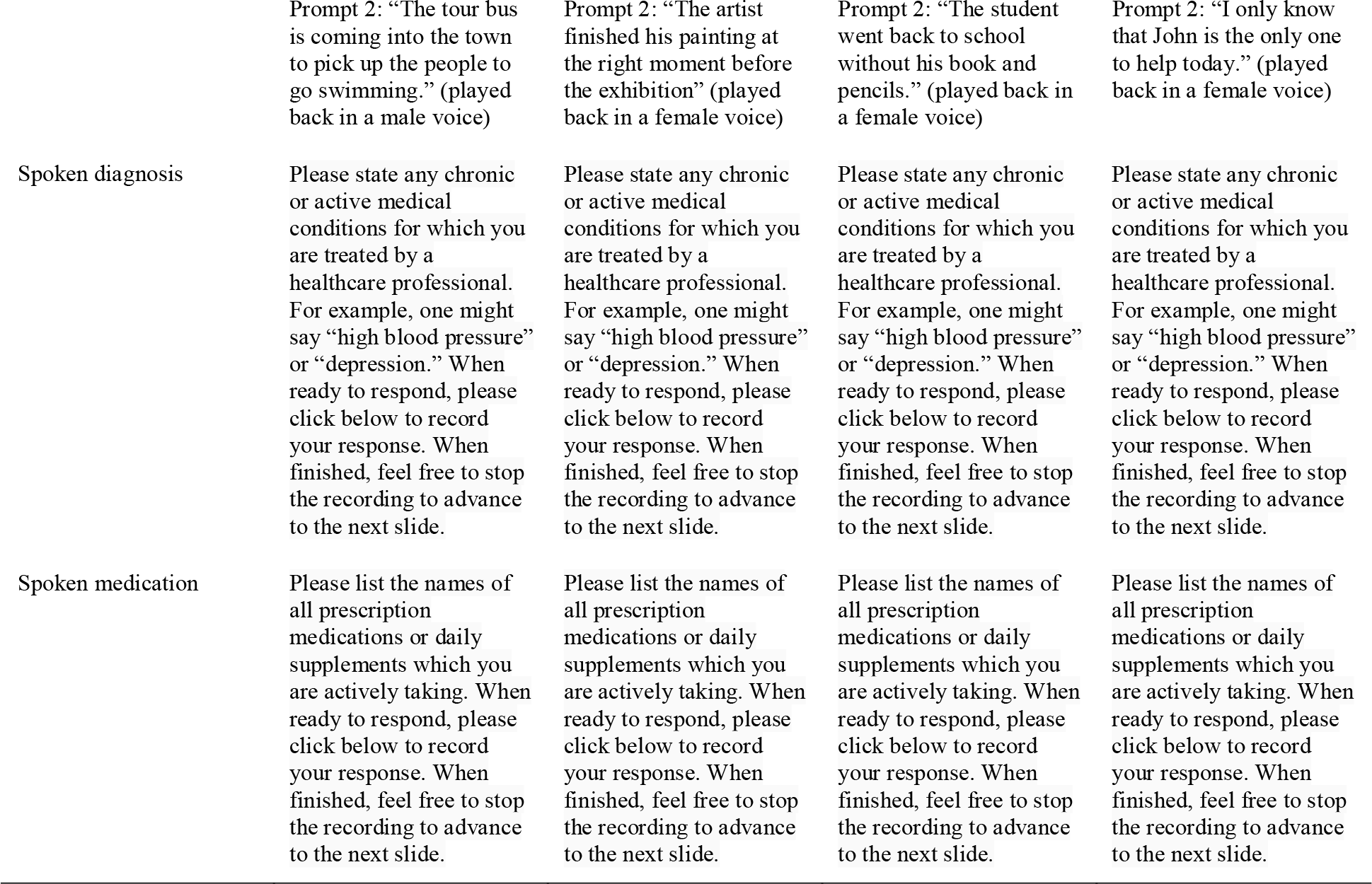
Voice prompt differences between survey versions A, B, C, and D. More details can be found on the Voiceome GitHub: https://github.com/jim-schwoebel/voiceome.

#### 1. Microphone test

Before participants could move on to the main part of the protocol, participants were asked to check whether their microphone was working. This check ensured that participant responses would be of a certain quality. The prompt was the following: “Please click the start button and then say: ‘The quick brown fox jumps over the lazy dog.’ You may press the Stop button if you finish before the timer runs out.” The reference string was ‘the quick brown fox jumps over the lazy dog.’ If a person repeated that phrase exactly, they would be given a score of 100% similarity.

#### 2. Free speech

Participants were asked to complete a single free speech task for 60 seconds. Four free speech tasks were used—one for each of the four different survey versions (Table 7). The prompts were, “Tell us about a recent happy memory based on experiences from the past month,” “Please list and briefly describe all the positive things that you expect to occur in the NEXT YEAR,” “Tell us about your hopes and dreams in what you plan to accomplish over the next THREE TO FIVE YEARS,” and “Describe the last moment that you remember when you were sad.” Participants were required to respond for the entire 60 seconds before they could move on to the next question. The free speech prompt was used to establish positive or negative valence (Cortes et al., 2021), which has been shown in multiple other studies to extract acoustic or linguistic information that may be relevant for conditions like depression and dementia (Sumali et al., 2020).

#### 3. Picture description

Participants were instructed to describe a picture that they saw on the screen for 60 seconds. Picture description tasks have been used to classify patients with Alzheimer’s disease symptoms versus age-matched controls (e.g., Forbes-McKay & Venneri, 2005). In the Voiceome Dataset, we used pictures of a man changing a lightbulb, a dog hiding after eating birthday cake, a cat and man stuck in a tree, and a family lounging at the beach (Table 7). Participants were required to respond for the entire 60 seconds before they could move on to the next question.

#### 4. Category naming

Category naming tasks are a common measure of semantic verbal fluency (e.g., Vaughan, Coen, Kenney, & Lawlor, 2018). This task asks participants to name all members of a category (e.g., animals) that they can think of as quickly as possible for a total of 60 seconds. This task has been used in previous literature to help monitor cognitive decline by counting of the total number of animals named in the time period, excluding repetitions, or the number of repetition or semantic errors (König et al., 2018). As before, participants re required to respond for the entire 60 seconds before they can move on to the next question. The four categories used—one for each survey version—were animals, tools, fruits, and household items.

A keyword dictionary was created in Python that included the top animal names (as nouns). A human reviewer examined this list for stopwords to exclude from analysis, and these stopwords were then discarded from analysis. The total number of correctly named animals were then represented as means and standard deviations.

#### 5. Phonemic fluency

Similarly to the category naming task, participants were asked to name all of the words they could think of that begin with a certain letter before 60 seconds passed. This task has been previously used to measure phonemic fluency and test memory in clinical study participants (e.g., Opasso, Barreto, & Ortiz, 2016). The four letters used in the Voiceome Protocol (one for each study version) were F, A, S, and H. Once again, participants are not able to stop the timer early and must speak for the entire 60 seconds.

A Python script was created that tokenized the transcript into words. The words that started with the letter F were summed for each session and were represented as means and standard deviations across all participants in the dataset.

#### 6. Phonetically-balanced paragraph reading

Reading passages has been used in previous research to measure the attention of participants (Feng, D’Mello, & Graesser, 2013). The four paragraphs used in the Voiceome Dataset—one for each survey version—included the Caterpillar passage (Patel et al., 2013), the grandfather passage (Darley, Aronson, & Brown, 1975), the rainbow passage (Fairbanks, 1960), and the North Wind and the Sun passage from Aesop’s Fables (Jesus, Valente, & Hall, 2015). These four passages are phonetically-balanced and the Caterpillar, grandfather, and rainbow passages have been used as standard protocol in other SLB-related studies.

#### 7. Sustained phonation

During this task, each participant is asked to say the vowel “/a/” for as long as they could hold their breath, with a maximum duration of 30 seconds (Maslan et al., 2011). This sustained phonation task has been used across a wide range of studies to measure motor symptoms, such as Parkinson’s disease (Wroge et al. 2018), as well as respiratory symptoms, such as COVID-19 (Cavallaro, Di Nicola, Quaranta, & Fiorella, 2021). The sustained phonation task also generalizes to individuals from various locations, accents, and languages. In this task, participants can stop the timer when they ran out of breath. The specific prompt used in the Voiceome Dataset was, “The goal of this task is to determine how long you can make the vowel sound ‘/a/ such as when one says the words ‘cheetah’ or ‘hallelujah.’ Click on the sample below to hear an example of the sound. When ready please start the recording, take a deep breath, and then say /a/ for as long as you can sustain the sound. Stop the recording when finished.”

#### 8. Diadochokinetic tasks

The Voiceome Protocol contains two diadochokinetic tasks, each of which have been used to measure psychomotor symptoms of muscles used in speech production. In the ‘pa-pa-pa’ task (Mahler, 2012), participants repeat the syllable “puh” (as in “possible” or “probable”) as many times as they can in 10 seconds. In the ‘pa-ta-ka’ task (Kaploun et al., 2011), participants repeat the syllables “puh,” “tuh,” and “kuh”, in that order, as quickly and accurately as they can in a 10-second window. These two tasks can help speech features measurements generalize across various regions, accents, and languages. The specific prompts are shown in Tables 5 and 6.

#### 9. Confrontational naming

In this task, a series of 25 images is displayed to the participant, who is asked to name each image within 10 seconds. For example, if an image that looks like a mushroom is displayed, a participant would be expected to say “mushroom.” Once they name the image, participants could click to view the next image. The number of correctly identified words (out of 25 images) is counted to quantify the ability of an individual to access and retrieve words as a means to identify anomia, aphasia, or cognitive decline (Fergadiotis, Hula, Swiderski, Lei, & Kellough, 2019). In the Voiceome Dataset, images were selected that match well onto the Boston Naming Test (Kaplan, Goodglass, & Weintraub, 1983), as demonstrated by Hall and colleagues (Hall, O’Carroll, & Frith, 2010). These images depicted a mixture of common objects (e.g., mushroom) and specific objects (e.g., corset). All images were presented in a black-and-white format. The 25 images used for each of the four versions of this task are noted in Table 7.

All 25 audio files per completed participant session were converted to mono 16,000 HZ using the SoX command line tool. After this, all these 25 audio files were combined into a single audio file for analysis, representing 1 master file with names images per completed session. This master file was then transcribed using DeepSpeech acoustic model version 0.7.0 (deepspeech-0.7.0-models.pbmm) combined with the language model (deepspeech-0.7.0-models.scorer). These transcripts were then analyzed with keyword frequency plots to determine the most common words used in all the naming tasks, in order to create a boundary of acceptable and unacceptable answers. This keyword acceptance list was used to automatically score how many images were properly named in the 25-image session. Participants who did not name more than 10 images were discarded from the analysis, or>40% correct was defined as a quality control criterion.

#### 10. Non-word pronunciation

Next, a series of ten pseudoword text strings appeared back-to-back on the screen. Participants were asked to pronounce each of the pseudowords within 10 seconds. Once they pronounced the word, they were able to move on to the next word. These pseudowords were of two types: those that have orthographically similar “neighbors” (e.g., plive→sounds like live) and those that have no neighbors (e.g., cogd). The pesudowords were selected based on the peer-reviewed literature, which has shown that patients with Alzheimer’s disease were mildly impaired relative to the healthy controls in reading pseudowords with neighbors, but were markedly impaired in reading pseudowords with no neighbors (Friedman 1992). The collection of non-words used in each of the four survey versions are detailed in Table 7.

All 10 non-word audio files per completed session were converted to mono 16,000 HZ using the SoX command line tool. After this, all these 10 audio files were combined into a single audio file for analysis, representing 1 master file with names images per completed session. This master file was then transcribed using DeepSpeech acoustic model version 0.7.0 (deepspeech-0.7.0-models.pbmm) combined with the language model (deepspeech-0.7.0-models.scorer). These transcripts were then analyzed with keyword frequency plots to determine the most common words used in all the naming tasks, in order to create a boundary of acceptable and unacceptable answers. This keyword acceptance list was used to automatically score how many images were properly named in the 10-non-word session. Participants that did not correctly name 4 or more non-words were discarded from the analysis, as they did not meet an *a priori* threshold for quality data.

#### 11. Memory recall

Finally, two sentence repeating tasks were used to test immediate memory recall ability, each of which lasted for 15 seconds. In this task, participants listened to a short audio passage, such as a speaker saying, *The man saw the boy that the dog chased*. The participants were then taken to a blank screen and were asked to repeat the sentence that they just heard. This task was then repeated with a separate prompt. In three of the four survey versions, both prompts were spoken by a female-sounding voice. In the other survey version, both prompts were spoken by a male-sounding voice. These prompts were created by our research team alongside expert neurologists to test immediate recall and were designed to replicate similar tasks used in clinical practice.

During this section of the study, the audio recordings of the participant began as soon as they saw the sentence—namely, before the participants were asked to recite the sentence with the blank screen in front of them. The recordings therefore not only capture the participants’ memory recall, but also all voice activity before they were asked to speak the required sentences. This pre-sentence recording information allows researchers to identify which type of device participants were using as speakers (e.g., headphones vs. loudspeakers). The speaker type can then be used to control for confounds in any statistical analysis of the data. Here, data from participants who were wearing headphones were discarded, whereas data from those who used laptop or phone speakers were kept for data analysis. This decision allowed the researchers to compare the transcripts to the playback recordings in order to check for errors. Specifically, by comparing the words heard by participants with the participant speech, false errors were minimized (e.g., if the audio was cut off and participants did not hear the whole phrase). Error rates for both tasks were taken together and averaged to compute a net score for immediate recall.

### Health-related questions

Participants were asked to speak responses to two optional health-related questions: (1) a list of all of their diagnosed health conditions and (2) a list all the medications that they were taking. The Microsoft Azure transcript was used for both analyses. Spoken diagnoses were put into a master list of strings and frequency distributions of keywords were extracted. A list of stopwords was assembled to remove common words (e.g., ‘the’ or ‘this’). After stopwords were removed, a frequency distribution of keywords was plotted using the Yellowbrick Python library.

In addition, a number of text-based survey questions were asked regarding health behaviors that may affect speech production. For example, the Voiceome Protocol includes single-item questions about a participant’s smoking history (one question for smoking frequency, one for smoking amount), diagnoses of high blood pressure or heart disease, previous surgeries around the head or neck area, the time of day that the participant woke up, the frequency with which they regularly exercise, whether or not the participant exercised before taking the survey, the number of hours slept the previous night, right or left-handedness, oral or dental problems, visual impairment, hearing impairment, and dyslexia. They were also asked whether they were suffering from the following conditions that day: cold, fever, shortness of breath, and cough.

10-point Likert scales were also used to assess how well participants felt while taking the survey, as well as stress, sleepiness, happiness, hydration, hunger, allergies, headache, pain, throat soreness, skin conditions, and overall quality of life. Furthermore, validated psychometric scales were used to measure a number of chronic and acute health conditions, such as the PHQ-9 (Kroenke, Spitzer, & Williams, 2001), the GAD-7 (Spitzer, Kroenke, Williams, & Löwe, 2006), a modified Altman Self-Rating Scale (Altman, Hedeker, Peterson, & Davis, 1997), The AUDIT-C questionnaire (Bush et al., 1998), A modified Sheehan Disability Scale (SDS; Sheehan, Harnett-Sheehan, & Raj, 1996), Part A of the ADHD Self-Report Scale (Kessler et al., 2005), the Insomnia Severity Index (Morin, 1993), and the Stanford Sleepiness Scale (Shahid, Wilkinson, Marcu, & Shapiro, 2011).

Finally, participants were asked to disclose certain demographic information, such as gender identity, age, level of education, employment status, marital status, total household income, fluency with the English language, height, and weight.

### Survey Interface

To enable data collection efforts, authors Jim Schwoebel and Austin New designed and built SurveyLex (https://surveylex.com), a web-enabled survey platform to create and distribute voice surveys. This product has been used by various research organizations to support a variety of SLB-related research studies and allows for voice surveys to be deployed as a URL link in the browser across a range of microphones and devices. Data was collected via a survey link and stored in cloud buckets encrypted on SurveyLex infrastructure.

All data collected from the Voiceome Dataset was downloaded using a command-line interface to a custom account on SurveyLex and was uploaded on S3 for later analysis by all authors. Data was exported, de-identified, and put into a password-protected S3 bucket with features and metadata for analysis.

### Study Protocol

The Voiceome Dataset was conducted from March 2019 through May 2020. All procedures were approved by the Western Institutional Review Board (WIRB), protocol WIRB® Protocol #20170781. Participants accessed the survey through an online link (https://voiceome.org). After signing the online informed consent form, participants who met eligibility criteria (described below) were routed to the main questionnaire. As depicted in Figure 7, the Voiceome Dataset was longitudinal in design, consisting of four main survey components.

Each session started with a microphone test prompt, in order to ensure that clinical study participants had access to a device compatible with the SurveyLex interface. Participants first completed a microphone test (as described in Task 1 above). This microphone test data was only used for testing the participant’s microphone before the start of each survey and the data was otherwise not analyzed. The first questionnaire (Survey A), was used to collect baseline speech measures, as well as to collect general information about the participants. This baseline questionnaire consisted of twelve types of speech tasks (in the order presented above in the Materials section), followed by questions relating to demographics and physical and mental health. This baseline survey protocol was designed to be able to be completed within 20-30 minutes and contained multiple breaks to minimize survey fatigue and hopefully lead to higher completion rates and higher-quality data.

Participants were asked to complete three follow-up survey(s) in the future, ideally each separated by one week. Accordingly, four versions of the main study were created: Survey A (the baseline questionnaire), Survey B, Survey C, and Survey D, all of which were designed to take 15-20 minutes. The differences among the four surveys are detailed in Table 7. Three speech tasks (sustained ‘/a/’ phonation, pa-pa-pa, pa-ta-ka) and all demographic and health questions were present in each of the four surveys (A-D). The remaining speech tasks varied among the four survey versions (detailed in the Materials section above and in Table 7), in order to test how various prompts affected utterances and whether learning occurred between various surveys. The question-order was not randomized in any of the four surveys in order to facilitate easier task switching and understanding by participants.

Participants were randomly assigned to one of four groups, each of which corresponded to an assignment of survey versions across the three longitudinal time slots after the baseline survey. The baseline survey was always Survey A. Group 1 (AAA) took Survey A in Weeks 2, 3, and 4. Group 2 (BAB) took Survey B in Weeks 2 and 4, but Survey A in Week 3. Group 3 (BCD) completed Survey B in Week 2, Survey C in Week 3, and Survey D in Week 4. Finally, Group 4 (CBD) completed Survey C in Week 2, Survey B in Week 3, and Survey D in Week 4.

### Audio Preprocessing

All speech recordings were converted to mono 16000 Hz wave files using the FFmpeg Python library. Acoustic features were extracted using OpenSMILE (Eyben, Wöllmer, & Schuller, 2010) and GeMAPS (Eyben et al., 2015), while linguistic features were extracted using the Allie repository (Schwoebel, 2020).

All speech text was transcribed using Microsoft Azure Speech to Text (https://azure.microsoft.com/en-us/services/cognitive-services/speech-to-text/). In addition to the Azure transcriptions, a subset of the audio files was also transcribed with additional automatic transcription services—Pocketsphinx (Huggins-Daines et al., 2006) and DeepSpeech version 0.7.0 (https://github.com/mozilla/DeepSpeech/releases/tag/v0.7.0)—and with crowd-sourced human transcription platforms (Rev.com, TranscribeMe). Rev.com and TranscribeMe were chosen as two external vendors that could manually transcribe audio files. Manual transcription was done in order to test the error rate of automated transcription techniques across a range of speech tasks (e.g., free speech, Caterpillar passage). Only speech recordings from participants who completed more than one Survey were transcribed.

## Data Availability

The four versions of the Voiceome Protocol can be found at the following SurveyLex links below. Researchers can easily clone these surveys for their own use by selecting the ‘templates’ feature during the SurveyLex survey design process.

- Survey A - https://app.surveylex.com/surveys/8a32cbb0-cc8a-11eb-9ea3-938cc8b6d71e
- Survey B - https://app.surveylex.com/surveys/061da3f0-a637-11eb-bcc9-eba67643f616
- Survey C - https://app.surveylex.com/surveys/a66494c0-a824-11ea-88c1-ab37bac1e1d4
- Survey D - https://app.surveylex.com/surveys/53737620-a637-11eb-bcc9-eba67643f616

To help with maximal replicability of this study design, all digital assets (audio, images, and text prompts) used for the trial were sourced either from open access Google searches, custom created by our research team, or acquired from other peer-reviewed articles. These are all available at https://github.com/jim-schwoebel/voiceome.

The complete data from the Voiceome Dataset can be accessed via a commercial license.

If you would like access to this data, please contact the corresponding author.

## Code Availability

Scripts used to generate the acoustic and linguistic features and reference ranges for this paper can be accessed at this link: https://github.com/jim-schwoebel/voiceome

This GitHub repository provides a convenient command line interface to reproduce our work and apply it in future research papers.

## Notes

### Competing Interest Statement

There is a conflict of interest.
Financial conflicts of interest exist for James Schwoebel, Lindsay A. Warrenburg, and Austin New as employees of Sonde Health Inc. (eg. equity ownership / stock options). Financial conflicts of interest also exist for Joel Schwartz, Roland Brown, Monroe Butler, Mark Moss, and Eleftheria K. Pissadaki who have been employed by Biogen Inc. and may have equity ownership, employee incentive plans, and/or stock options in Biogen Inc.

### Funding Statement

Sonde Health Inc. and Biogen Inc. provided funding for this work. Financial conflicts of interest exist for James Schwoebel, Lindsay A. Warrenburg, and Austin New as employees of Sonde Health Inc. (eg. equity ownership / stock options). Financial conflicts of interest also exist for Joel Schwartz, Roland Brown, Monroe Butler, Mark Moss, and Eleftheria K. Pissadaki who have been employed by Biogen Inc. and may have equity ownership, employee incentive plans, and/or stock options in Biogen Inc.

### Author Declarations

All procedures were approved by the Western Institutional Review Board (WIRB), protocol #20170781.

